# Prevalence and Factors associated with Mental health impact of COVID-19 Pandemic in Bangladesh: A survey based cross sectional study

**DOI:** 10.1101/2021.01.05.21249216

**Authors:** Tanvir Abir, Nazmul Ahsan Kalimullah, Uchechukwu L Osuagwu, Dewan Muhammad Nur –A Yazdani, Taha Husain, Piwuna Christopher Goson, Palash Basak, Md Adnan Rahman, Abdullah Al Mamun, P. Yukthamarani Permarupan, Md Yusuf Hossein Khan, Kingsley Agho

**Affiliations:** Associate Professor, College of Business Administration—CBA, International University of Business, Agriculture and Technology—IUBAT University, Dhaka 1230, Bangladesh; Vice-chancellor, Begum Rokeya University, Rangpur-5404, Bangladesh; Diabetes, Obesity and Metabolism Translational Research Unit, Western Sydney University, Campbelltown, NSW 2560,Australia; Assistant Professor, College of Business Administration—CBA, International University of Business, Agriculture and Technology—IUBAT University, Dhaka 1230, Bangladesh; Lecturer, Department of Gender and Development Studies, Begum Rokeya University, Rangpur 5404, Bangladesh; Department of Psychiatry, College of Health Sciences, University of Jos, Nigeria; School of Environment and Life Sciences (Environmental Science and Management), University of Newcastle, Callaghan 2308,Australia; Senior lecturer, College of Business Administration (CBA), International University of Business Agriculture and Technology (IUBAT), Dhaka-1230, Bangladesh; Faculty of Business and Management, UCSI University, Kuala Lumpur 56000, Malaysia; Faculty of Entrepreneurship and Business, Universiti Malaysia Kelantan, Kota Bharu 16100, Malaysia; College of Tourism and Hospitality Management IUBAT—International University of Business Agriculture and Technology, Dhaka, Bangladesh, PhD Researcher in Tourism University of Algarve, Faro Portugal; School of Health Science, Western Sydney University, Campbelltown, NSW 2560, Australia.; African Vision Research Institute (AVRI), University of KwaZulu-Natal, Westville Campus, Durban, 3629, South Africa

**Keywords:** mental health, depression, anxiety, stress, coronavirus, divisions, Bangladesh

## Abstract

**Background:** Feelings of isolation, insecurity, and instability triggered by COVID-19 could have a long-term impact on the mental health status of individuals. This study examined the prevalence and factors associated with the mental health symptoms of anxiety, depression, and stress during the COVID-19 pandemic in Bangladesh.

**Methods:** From 1^st^ – 30^th^ April 2020, we used a validated self-administered questionnaire to conduct a cross-sectional study on 10,609 participants through an online survey platform. We assessed mental health status using the Depression, Anxiety, and Stress Scale (DASS-21). The total depression, anxiety, and stress subscale scores were divided into normal, mild, moderate, severe, and multinomial logistic regression was used to examine associated factors.

**Results:** The prevalence of depressive symptoms was 15%, 34%, and 15% for mild, moderate, and severe depressive symptoms, respectively. The prevalence of anxiety symptoms was 59% for severe anxiety symptoms, 14% for moderate anxiety symptoms, and 14% for mild anxiety symptoms while, the prevalence for stress levels were 16% for severe stress level, 22% for moderate stress level and 13% for mild stress level. Multivariate analyses revealed that the most consistent factors associated with mild, moderate, and severe of the three mental health subscales (depression, anxiety, and stress) were respondents who lived in Dhaka and Rangpur division, females, those who self-quarantine in the previous 7 days before the survey and those respondents who experienced chills, breathing difficulty, dizziness, and sore throat.

**Conclusion:** Our results showed that about 64%, 87%, and 61% experienced depressive symptoms, anxiety symptoms, and levels of stress, respectively. In Bangladesh, there is a need for better mental health support for females especially those that lived in Dhaka and Rangpur division and experienced chills, breathing difficulty, dizziness, and sore throat during COVID-19 and other future pandemics.

## Introduction

As the global population tries to make sense of the transformations including personal adjustments to lifestyle brought about by the COVID-19 pandemic, residents of low to middle-income countries (LMIC) including Bangladesh face greater challenges due to the fragile health systems [**1, 2**], the dense population of Bangladesh and the fact that the country houses a million stateless Rohingya refugees in sprawling refugee camps that are conducive to the spread of epidemics. Bangladesh also has significant migrant populations living in Italy, a COVID-affected country [1]. Whilst the mortality rates in Bangladesh have remained low, due to the timing of the infection, the early transmission of the virus, and the response to the pandemic by authorities, the low socio-economic status of the country and the existing health inequalities usually lead to worse effects [**3**].

Science has played a significant role in improving people’s understanding of the virus, finding effective ways of containment through timely sequencing of the virus and rapid sharing of the data [**4**], and most recently the development of different vaccines. Unraveling the genetic sequence of the SARS-COV-2 virus about 4 weeks after the outbreak of the SARS-COV-2 virus [**5**], which was shorter compared to the Spanish flu which took almost seven decades for the scientist to unravel the genetic sequence of the disease [**6**], which is crucial to the development of a diagnostic test and potential treatment [**7, 8**].

Globally, the virus has infected over 84 million people, including 1.8 million reported deaths from the infection. In Bangladesh, there have been 515,000 confirmed cases as of 2^nd^ January 2021, with 7576 deaths reported to the WHO [**9**]. The COVID-19 pandemic spread faster, and the mortality rate is higher than those attributed to the Middle Eastern respiratory syndrome (MERS) and Severe acute respiratory syndrome (SARS). Thus, there was fear and panic among residents as news about its fatal nature spread very easily through traditional and social media outlets, leaving behind a trail of despair and disruptions in lifestyle. The high mortality rate, closure of businesses, and strict containment measures by national governments also added to the hidden and unhidden mental health burden of the pandemic [**10, 11**]. However, no one has been able to report comprehensively the mental health impact of the pandemic in most Low and Middle Income Countries (LMIC).

Despite the delay in COVID 19 cases in Bangladesh (the first case reported 18^th^ March 2020), the country’s global supply chain of International fashion brands and human resource exports suffered a huge set-back with devastating psychosocial consequences emanating from the International and local economic impacts [**12**]. The rapid spread of the infection and the business climate in Bangladesh, caused fear, worry, and stress as restrictions were put in place by the government. Post-traumatic stress symptoms as well as delayed grief and the sense of loss after multiple deaths and loss of jobs and avenue to socialize have been reported in previous studies [**13**].

Studies have reported that the COVID-19 pandemic has significant negative impacts on the mental health of college students, with female students reporting higher levels of perceived stress and inability to focus on their academic work [**14**]. Similarly, couples undergoing Assisted Reproductive Treatment showed the severe psychological impact of the pandemic particularly among women who were more emotionally distressed, anxious, and depressed than their men counterparts. This is because Assisted Reproductive Treatment was stopped in many centers due to rising concern on the impact of COVID-19 on pregnancy [**15**]. Coupled with these, are the uncertainties about effective treatment, availability of effective vaccine as well as whether life could return to normal.

All these could negatively affect the mental health of the populace and by extension, the productivity of the country that depends largely on international trading, which has so far been decimated by the pandemic. Health care delivery in Bangladesh has major challenges including weak governance and over-centralized framework, the fact that over 58% of all the physicians work in the private sector that is poorly regulated, and the lack of funding for the public sector [**16**]. Layered on top of these, are the lack of resources and disproportionate distribution of mental health services in Bangladesh, leading to poor access to mental health facilities and care [**17**]. Therefore, this study aims to determine the prevalence and factors associated with mental health symptoms in the population, which could drive the people’s economy and wellbeing during and after the pandemic.

## Methods

### Study Design

This cross-sectional online study was conducted from April 1-30, 2020 corresponding to the mandatory lockdown period imposed in different parts of Bangladesh. The Institutional Review Board of Dr. Wazed Research and Training Institute, Begum Rokeya University, Rangpur (#BRUR/DWRTI/a.n.004), approved the study. This study follows the tenets of the Declaration of Helsinki as revised in Fortaleza. Informed consent was obtained through an online preamble before the respondents began the questionnaire. The participants were assured of the confidentiality of the information provided and their freedom of choice of participation.

A self-administered survey was created using a google form, and since traditional face-to-face interviews were not possible due to the lockdown, social media platforms (e.g. Facebook, Google Plus, and Google Form) were used for the distribution of the survey to reach the target respondents living in different parts of Bangladesh. The respondents clicked the link on the platform and responded to the survey voluntarily. This was a convenient sample, and the survey was anonymous, and confidentiality of data was ensured.

### Study Population

The respondents had to be resident in Bangladesh and able to provide online informed consent. To be eligible for participation, participants had to be 18 years and over, and should be able to provide online consent.

### Measurements and Covariates

The covariates, which are shown in Table 1, were classified into four parts: demographic, household factors, compliance with health measures, and health-related factors. The first part gathered demographic information of the participants, including gender, age, living area (division), level of education, marital status, and working status. The second part was the household factors, which asked about living arrangements, the number living together, and the third part included COVID-19 factors, which asked whether the participants have been tested for COVID-19. The fourth part evaluated the compliance to WHO recommended precautionary measures, which included avoiding crowded gatherings, handshaking and use of public transport, wear of facemask when going out, advocate other people about the health risk of the infection. The fifth part evaluated the history of health-related symptoms (if the respondents had experienced any of the following symptoms: fever, pain, headache, chills persistent, dizziness, and breathing difficulties, a couple of weeks before data collection). The information about these five parts is listed in Supplementary Table 1.

**Table 1.**
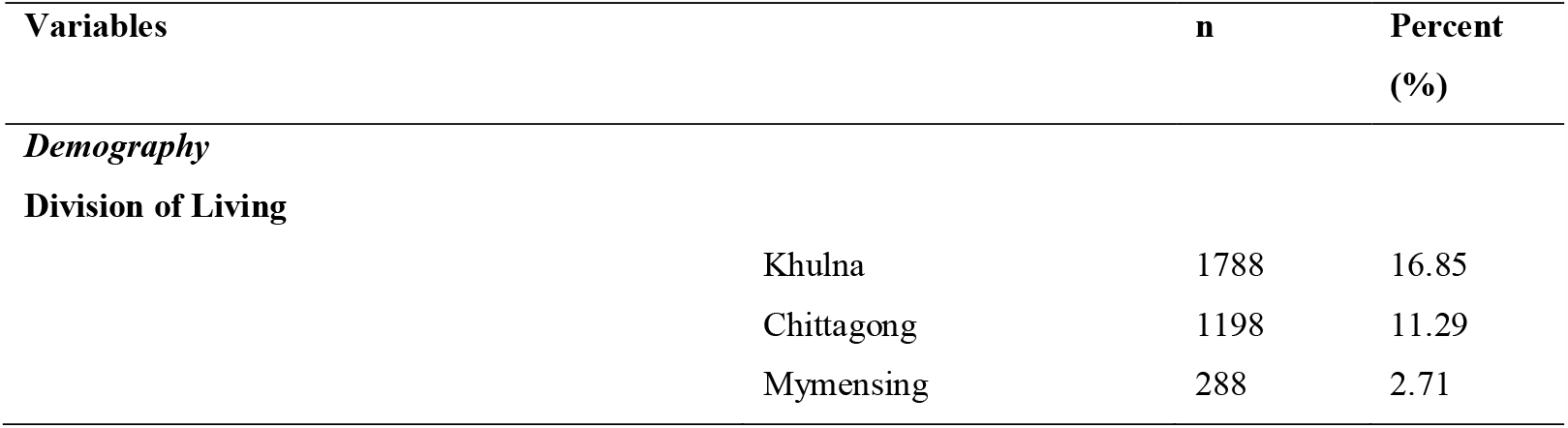
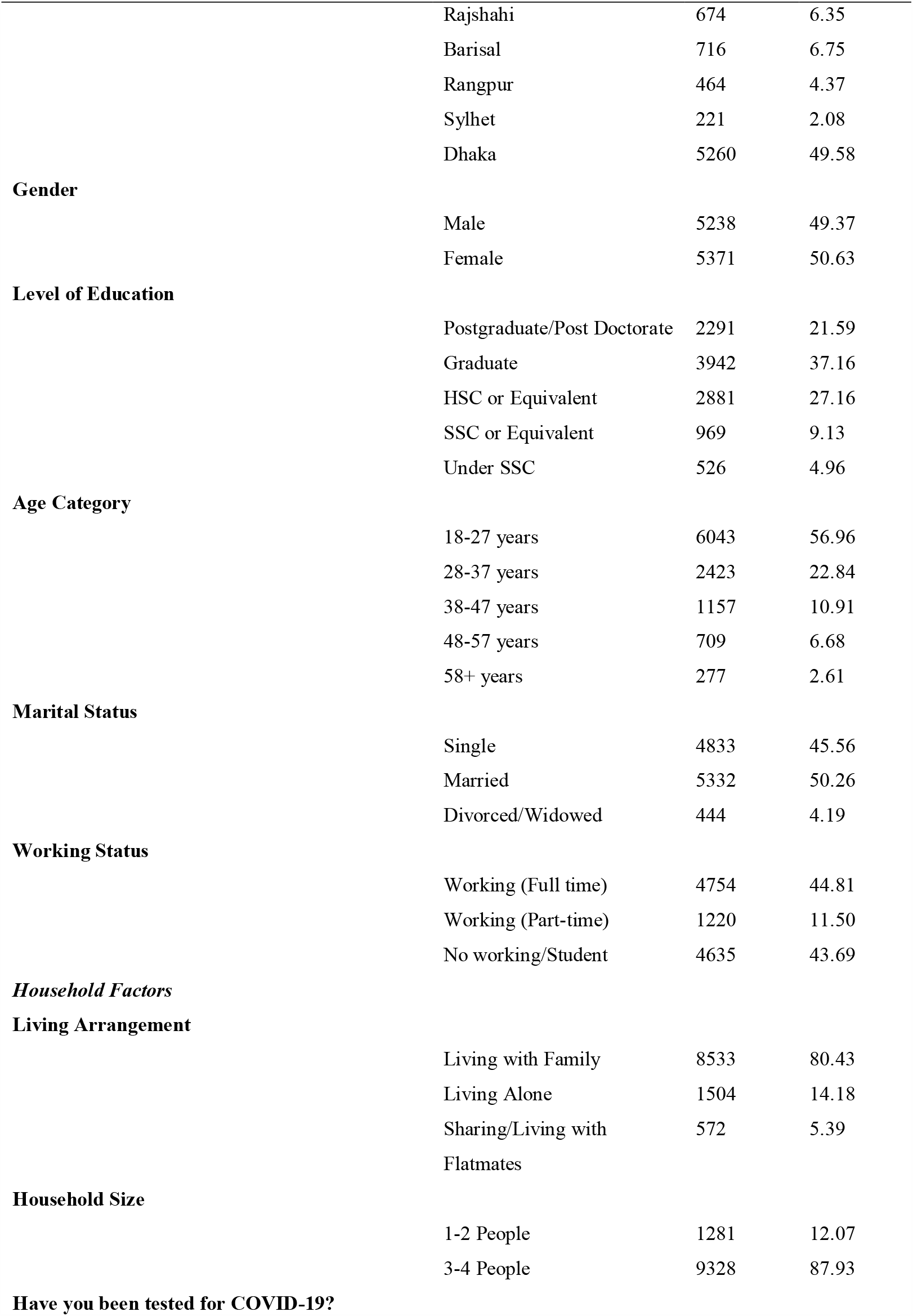
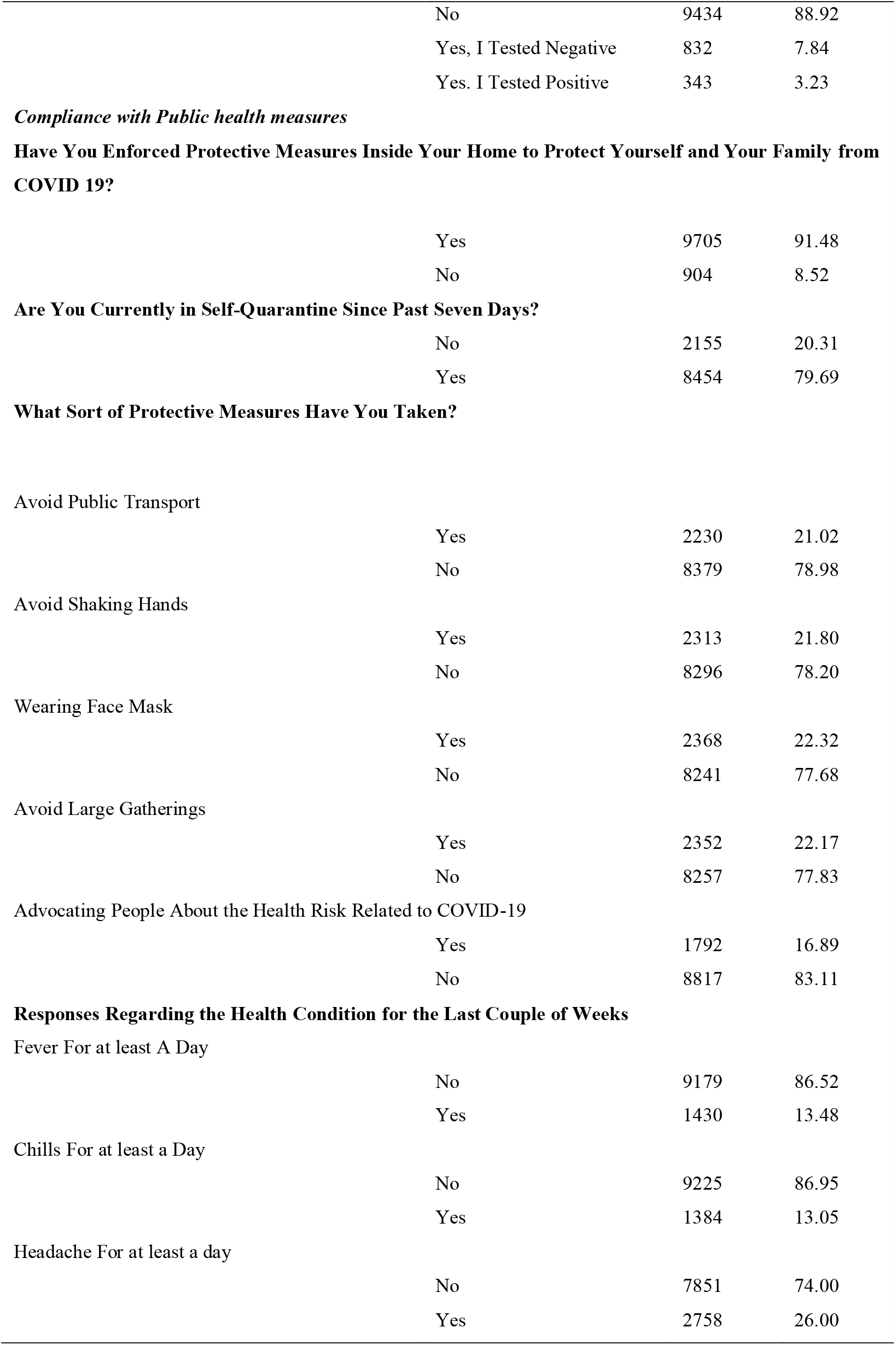
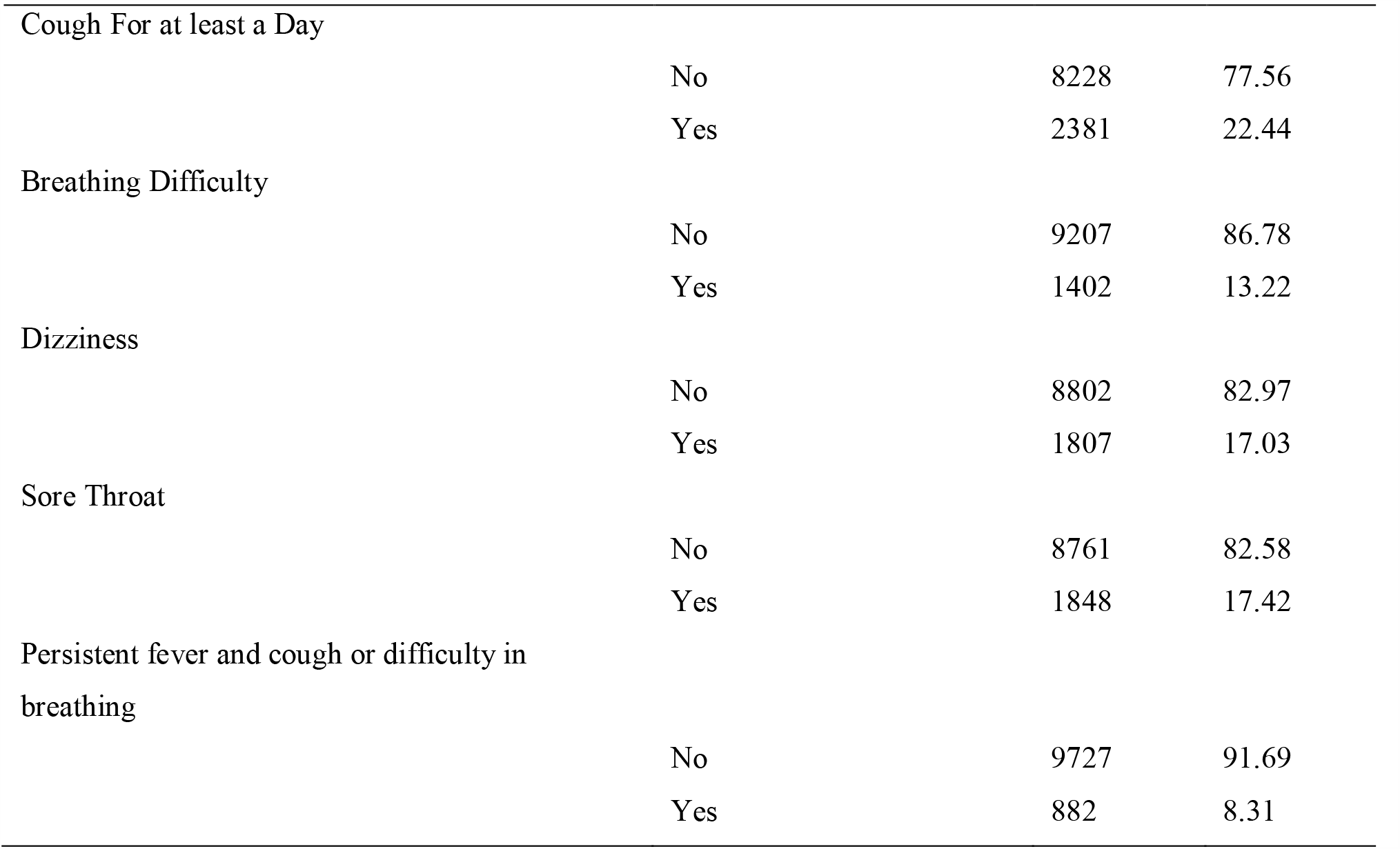
Characteristics of the study population (n=10609)

| Variables | n | Percent (%) |
| --- | --- | --- |
| *Demography* |  |  |
| **Division of Living** |  |  |
| Khulna | 1788 | 16.85 |
| Chittagong | 1198 | 11.29 |
| Mymensing | 288 | 2.71 |

|  |  |  |  |
| --- | --- | --- | --- |
|  | Rajshahi | 674 | 6.35 |
|  | Barisal | 716 | 6.75 |
|  | Rangpur | 464 | 4.37 |
|  | Sylhet | 221 | 2.08 |
|  | Dhaka | 5260 | 49.58 |
| **Gender** |  |  |  |
|  | Male | 5238 | 49.37 |
|  | Female | 5371 | 50.63 |
| **Level of Education** |  |  |  |
|  | Postgraduate/Post Doctorate | 2291 | 21.59 |
|  | Graduate | 3942 | 37.16 |
|  | HSC or Equivalent | 2881 | 27.16 |
|  | SSC or Equivalent | 969 | 9.13 |
|  | Under SSC | 526 | 4.96 |
| **Age Category** |  |  |  |
|  | 18-27 years | 6043 | 56.96 |
|  | 28-37 years | 2423 | 22.84 |
|  | 38-47 years | 1157 | 10.91 |
|  | 48-57 years | 709 | 6.68 |
|  | 58+ years | 277 | 2.61 |
| **Marital Status** |  |  |  |
|  | Single | 4833 | 45.56 |
|  | Married | 5332 | 50.26 |
|  | Divorced/Widowed | 444 | 4.19 |
| **Working Status** |  |  |  |
|  | Working (Full time) | 4754 | 44.81 |
|  | Working (Part-time) | 1220 | 11.50 |
|  | No working/Student | 4635 | 43.69 |
| ***Household Factors*** |  |  |  |
| **Living Arrangement** |  |  |  |
|  | Living with Family | 8533 | 80.43 |
|  | Living Alone | 1504 | 14.18 |
|  | Sharing/Living with Flatmates | 572 | 5.39 |
| **Household Size** |  |  |  |
|  | 1-2 People | 1281 | 12.07 |
|  | 3-4 People | 9328 | 87.93 |
| **Have you been tested for COVID-19?** |  |  |  |

|  |  |  |  |
| --- | --- | --- | --- |
|  | No | 9434 | 88.92 |
|  | Yes, I Tested Negative | 832 | 7.84 |
|  | Yes. I Tested Positive | 343 | 3.23 |
| ***Compliance with Public health measures*** |  |  |  |
| **Have You Enforced Protective Measures Inside Your Home to Protect Yourself and Your Family from COVID 19?** |  |  |  |
|  | Yes | 9705 | 91.48 |
|  | No | 904 | 8.52 |
| **Are You Currently in Self-Quarantine Since Past Seven Days?** |  |  |  |
|  | No | 2155 | 20.31 |
|  | Yes | 8454 | 79.69 |
| **What Sort of Protective Measures Have You Taken?** |  |  |  |
| Avoid Public Transport |  |  |  |
|  | Yes | 2230 | 21.02 |
|  | No | 8379 | 78.98 |
| Avoid Shaking Hands |  |  |  |
|  | Yes | 2313 | 21.80 |
|  | No | 8296 | 78.20 |
| Wearing Face Mask |  |  |  |
|  | Yes | 2368 | 22.32 |
|  | No | 8241 | 77.68 |
| Avoid Large Gatherings |  |  |  |
|  | Yes | 2352 | 22.17 |
|  | No | 8257 | 77.83 |
| Advocating People About the Health Risk Related to COVID-19 |  |  |  |
|  | Yes | 1792 | 16.89 |
|  | No | 8817 | 83.11 |
| **Responses Regarding the Health Condition for the Last Couple of Weeks** |  |  |  |
| Fever For at least A Day |  |  |  |
|  | No | 9179 | 86.52 |
|  | Yes | 1430 | 13.48 |
| Chills For at least a Day |  |  |  |
|  | No | 9225 | 86.95 |
|  | Yes | 1384 | 13.05 |
| Headache For at least a day |  |  |  |
|  | No | 7851 | 74.00 |
|  | Yes | 2758 | 26.00 |
| Cough For at least a Day |  |  |  |
|  | No | 8228 | 77.56 |
|  | Yes | 2381 | 22.44 |
| Breathing Difficulty |  |  |  |
|  | No | 9207 | 86.78 |
|  | Yes | 1402 | 13.22 |
| Dizziness |  |  |  |
|  | No | 8802 | 82.97 |
|  | Yes | 1807 | 17.03 |
| Sore Throat |  |  |  |
|  | No | 8761 | 82.58 |
|  | Yes | 1848 | 17.42 |
| Persistent fever and cough or difficulty in breathing |  |  |  |
|  | No | 9727 | 91.69 |
|  | Yes | 882 | 8.31 |

The Depression, Anxiety and Stress Scale - 21 Items (DASS-21) is a set of three self-report scales designed to measure the emotional states of depression, anxiety, and stress and were calculated based on a previous study [**18**]. Responses to each item were rated from 0 to 3, where “never” (score ‘0’) to ‘almost always” (score ‘3’). Item 3, 5, 10, 13, 16, 17, and 21 (see supplementary table 1) were classified as the depression subscale, and the total depression subscale score was multiplied by 2 to calculate the final score and divided into normal (0–9), mild depression (10–13), moderate depression (14–20), severe depression (21+). The Cronbach’s alpha coefficients measuring internal consistency among the depression subscale score ranged from 0.75 to 0.77, indicating a satisfactory level of reliability.

The items 2, 4, 7, 9, 15, 19, and 20 (see supplementary table 1) formed the anxiety subscale, and the total anxiety subscale score was multiplied by 2 to calculate the final score and divided into normal (0–7), mild anxiety (8-9), moderate anxiety (10–14), severe anxiety (15+) [**18**]. The Cronbach’s alpha coefficient for anxiety subscale scores ranged from 0.65 to 0.77, indicating acceptable internal consistency.

Items 1, 6, 8, 11, 12, 14, and 18 (see supplementary table 1) were classified as the stress subscale. The total stress subscale score was multiplied by 2 to calculate the final score and was divided into normal (0–14), mild stress (15–18), moderate stress (19–25), severe stress (26+), and extremely severe stress (35–42) [**18**]. The Cronbach’s alpha coefficient of stress subscale scores ranged from 0.78 to 0.81, indicating an acceptable level of internal consistency.

### Statistical Analysis

Descriptive statistics using frequency tabulations were used to present the sample characteristics. The prevalence and 95% confidence intervals of symptoms of depression, anxiety, and levels of stress were calculated for normal, mild, moderate, and severe. The association was further tested by odds ratios (OR) using univariate multinomial logistic regression, and multiple multinomial logistic regression analyses were performed to identify significant factors associated with symptoms of depression, anxiety, and levels of stress.

In the multiple multinomial logistic regression analyses, four-stage modeling was employed. In the first stage, the demographic factors were entered into the first stage model. We conducted a manually executed elimination method to determine factors associated with symptoms of depression, anxiety, and levels of stress at (p <0.05). The significant factors in the first stage were added to the household factors in the second stage model; this was then followed by the elimination procedure. We used a similar statistical approach for compliance with public health and Health condition\factors in the third and fourth stages, respectively. Associations were presented as unadjusted OR (95% CI) for all explanatory variables and then adjusted OR (95% CI) for the variables retained in the final step. All statistical analyses were conducted using STATA/MP Version.14.1 (Stata Corp, College Station, TX, USA).

### Spatial Analysis

We conducted spatial distribution for mental health which consisted of depression, anxiety, and stress subscales. A series of maps were prepared using ArcGIS Desktop 10.8 [19]. The average level of depression, anxiety, and stress for the first-level administrative unit of Bangladesh (division) was calculated based on the identification of factors through map comparisons and regression analysis for mild, moderate, and severe levels. In the maps, the adjusted odds ratios (AOR) for each level was categorized into five quantiles and were presented using graduated colour symbols.

## Results

### Demographic Characteristics

From the 10,900 participants, data for 10,660 respondents (aged 18 years and over) deriving out of all 8 divisions in Bangladesh were included in the analysis. 5238 participants (49.1%) were males and mostly aged between 18-37years (8466, 79.4%). Of the total number of respondents, 6233 (58.5%) had a university degree or higher, and 5332 (50.3%) were married. About half of the participants (5260 participants [49.6%]) lived in Dhaka division, with the family (8533, 80.4%) or with up to 3 people (9328, 87.9%) and were working full time or part-time (5974 participants [56.0%]) during the pandemic. This survey included data from 343 individuals (3.2%) with confirmed cases of COVID-19 and 832 individuals (7.8%) with suspected cases of COVID-19.

Of the total number of respondents, the majority enforced protective measures in their homes to protect their families (9705, 91.5%), had quarantine experience, 8454 (79.7%) but more than two-third (∼78% each) did not comply with the public health measures of avoiding public transport, handshaking, large gatherings, wear of facemask when going out, which were put in place to contain the spread of the disease. More than two-thirds of the participants had at least one symptom of COVID-19 a couple of weeks before data collection, especially persistent fever and cough (9727, 91.7%) and difficulty breathing (9207, 86.8%). Additional demographic and epidemic-related characteristics are presented in Table 1.

### Prevalence and 95% confidence intervals (CI) of Covid-19 depression, anxiety, and stress

The prevalence of symptoms for the 3 mental health conditions among the total sample is shown in Figures 1 (a-c) for depression, anxiety, and stress, respectively. The prevalence was 65.0% for depression (6822 participants total, including 1563 participants [14.7%, 95%CI, 14.0-15.4%] with mild depression and 1552 participants [14.6%, 95%CI, 13.9-15.3%] with severe depression), 86.9% for anxiety (9267 participants total, including 1440 participants [13.5%, 95%CI, 12.4-13.7%] with mild anxiety and 6310 participants [59.2%, 95%CI, 58.3-60.1%] with severe anxiety), 50.5% (95% CI, 28.8%-29.6%) for stress (5381 participants total, including 1339 participants [12.6%, 95%CI, 12.0-13.2%] with mild stress and 1684 participants [15.8%, 95% CI, 15.1-16.5%] with severe stress). Additional details on the prevalence of mental health symptoms in different populations are presented in Figure 1.

**Figure 1 (a):**
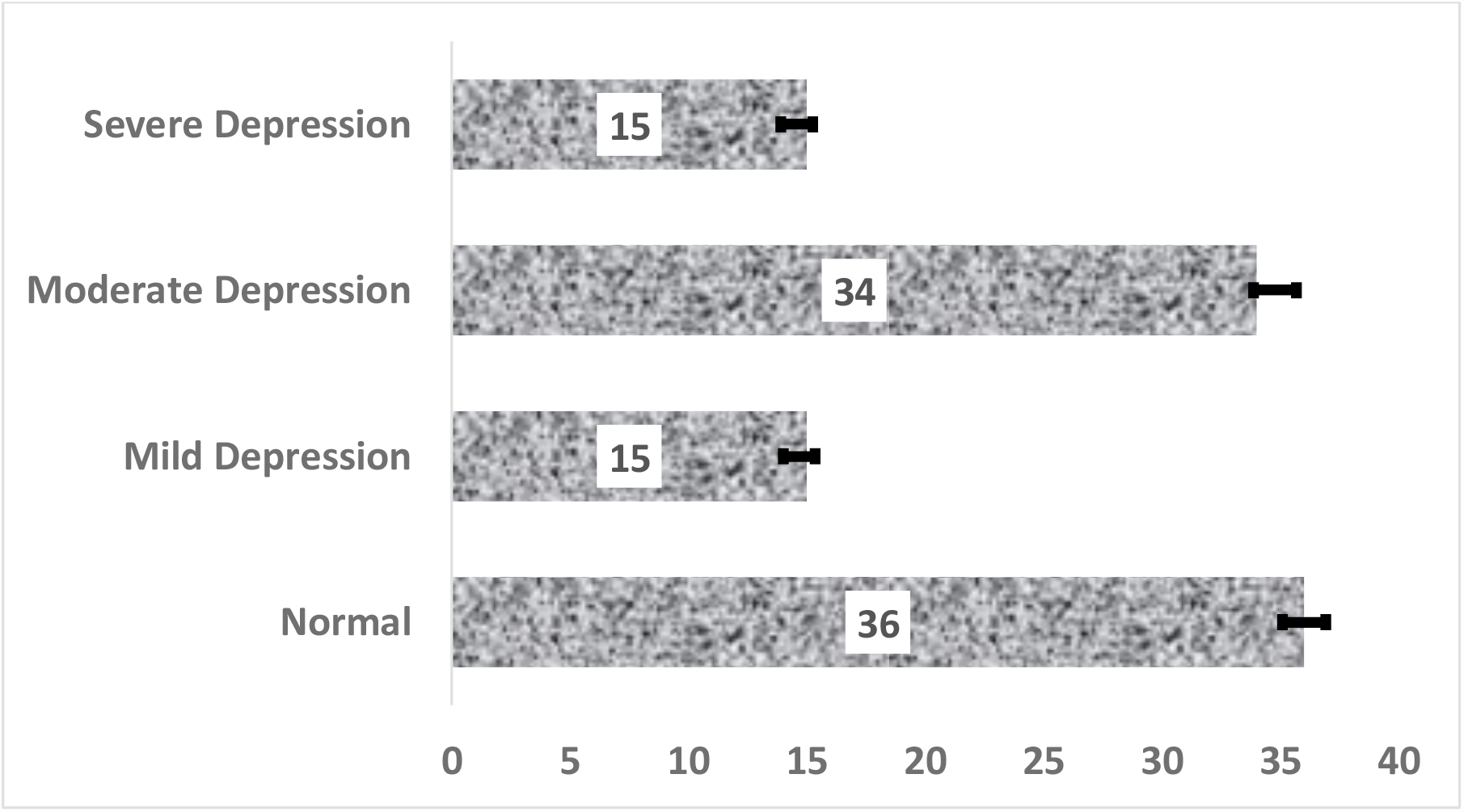
Prevalence and 95% CI of Depression level during COVID-19

**Figure 1 (b):**
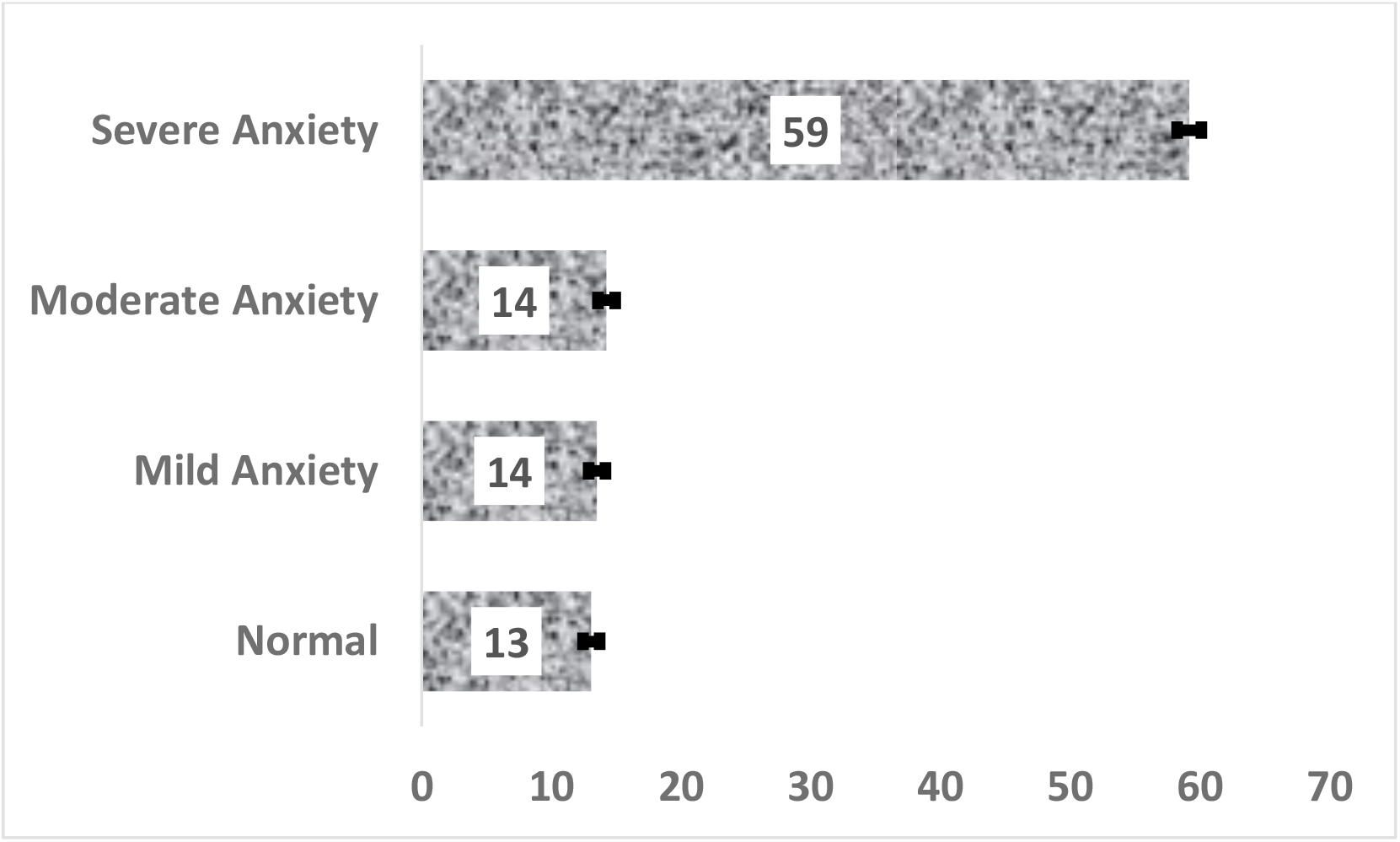
Prevalence and 95% CI of Anxiety level during COVID-19

**Figure 1 (c):**
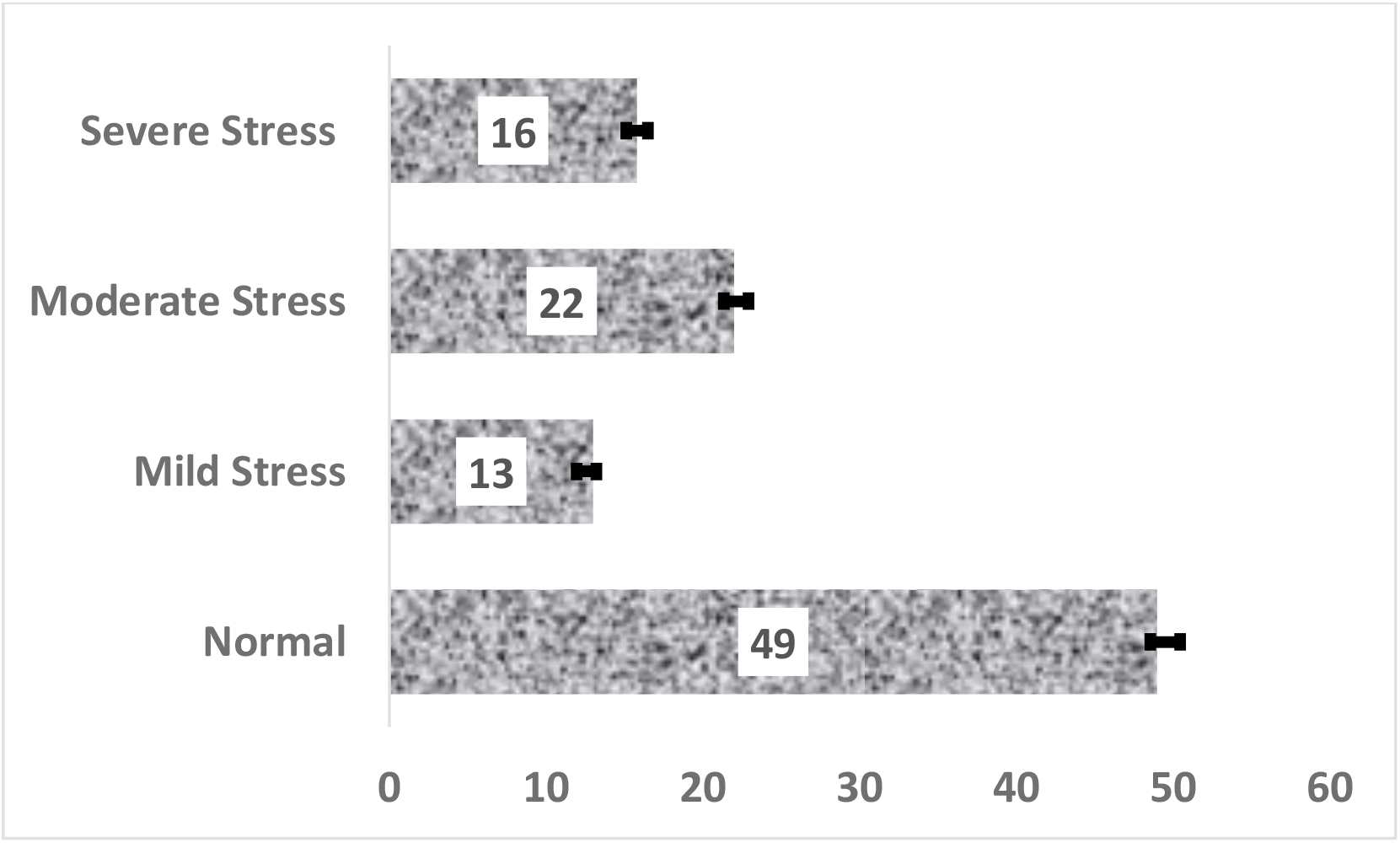
Prevalence and 95% CI of Stress level during COVID-19

### Factors Associated with Symptoms of Depression, Anxiety, and Stress

The results of the unadjusted analysis of demographic and COVID-19 related variables are presented in the supplementary Tables 1-3, for depression, anxiety, and stress, respectively. Table 2 presents the multivariate analysis of factors associated with depression in this cohort. In the multivariable analysis, being married, having lower than a postgraduate degree, living alone or living in shared accommodation, living in the Sylhet, Dhaka, Chittagong, Rajshahi, and, Rangpur divisions, having experienced any COVID-19 related health symptom couple of weeks before data collection, were found to be associated with symptoms of depression at all levels. Figure 2 presents the adjusted odds ratios for depression symptoms among the respondents by region. The map in Figure 2, indicates that significantly high odds of depression occurred in all 8 regions of Bangladesh with the highest odds of severe depression reported in Rangpur, Sylhet and Chittagong reported (darkest brown colour). Individuals who were tested for COVID-19 also had remarkably higher levels of mild-severe symptoms of depression compared to those that had not been tested. In addition, female participants, those who were divorced or separated, residents of Barisal or Mymensing divisions, and those who traveled by public transport displayed a higher risk of moderate-severe symptoms of depression. Additional details on the factors associated with depression are presented in Table 2. Individuals with confirmed COVID-19 had at least a 50 percent higher risk of moderate-severe depression symptoms compared to those who were not tested for the disease.

**Table 2:** Factors associated with mild, moderate, and severe depression during COVID-19 in Bangladesh.

| Variables | Mild |  | Moderate |  | Severe |  |
| --- | --- | --- | --- | --- | --- | --- |
|  | aOR | Pvalue | aOR | Pvalue | aOR | Pvalue |
| **Demography** |  |  |  |  |  |  |
| **Division of Living** |  |  |  |  |  |  |
| Khulna | **1.00** |  | **1.00** |  | **1.00** |  |
| Chittagong | 2.16 (1.65, 2.81) | <0.001 | 4.38 (3.47, 5.55) | <0.001 | 6.38 (4.77, 8.56) | <0.001 |
| Mymensing | 0.96 (0.59, 1.57) | 0.875 | 2.68 (1.86, 3.85) | <0.001 | 2.51 (1.57, 4.01) | <0.001 |
| Rajshahi | 1.42 (1.01, 1.98) | 0.042 | 3.56 (2.74, 4.62) | <0.001 | 3.77 (2.70, 5.27) | <0.001 |
| Barisal | 1.04 (0.77, 1.41) | 0.782 | 1.76 (1.36, 2.27) | <0.001 | 2.09 (1.50, 2.93) | <0.001 |
| Rangpur | 1.91 (1.28, 2.86) | 0.002 | 5.07 (3.65, 7.03) | <0.001 | 5.68 (3.82, 8.45) | <0.001 |
| Sylhet | 5.34 (2.93, 9.75) | <0.001 | 10.67 (6.05, 18.81) | <0.001 | 15.17 (8.07, 28.53) | <0.001 |
| Dhaka | 1.85 (1.54, 2.22) | <0.001 | 2.48 (2.11, 2.93) | <0.001 | 2.88 (2.30, 3.62) | <0.001 |
| **Gender** |  |  |  |  |  |  |
| Male | **1.00** |  | **1.00** |  | **1.00** |  |
| Female | 1.14 (0.99, 1.30) | 0.060 | 1.63 (1.45, 1.83) | <0.001 | 1.71 (1.47, 1.98) | <0.001 |
| **Level of Education** |  |  |  |  |  |  |
| Postgraduate/Post Doctorate | **1.00** |  | **1.00** |  | **1.00** |  |
| Graduate | 1.47 (1.24, 1.75) | <0.001 | 1.64 (1.41, 1.90) | <0.001 | 1.55 (1.28, 1.88) | <0.001 |
| HSC or Equivalent | 1.25 (1.03, 1.52) | 0.026 | 2.00 (1.70, 2.35) | <0.001 | 2.19 (1.79, 2.68) | <0.001 |
| SSC or Equivalent | 1.53 (1.14, 2.05) | 0.005 | 3.57 (2.84, 4.48) | <0.001 | 3.22 (2.44, 4.24) | <0.001 |
| Under SSC | 1.61 (1.14, 2.28) | 0.007 | 2.64 (1.97, 3.53) | <0.001 | 2.89 (2.04, 4.10) | <0.001 |
| **Age in category** |  |  |  |  |  |  |
| 18-27 years | **1.00** |  | **1.00** |  | **1.00** |  |
| 28-37 years | 1.16 (0.94, 1.44) | 0.166 | 1.21 (1.02, 1.43) | 0.032 | 1.49 (1.23, 1.82) | <0.001 |
| 38-47 years | 0.84 (0.63, 1.13) | 0.246 | 1.10 (0.89, 1.37) | 0.367 | 0.80 (0.61, 1.04) | 0.090 |
| 48-57 years | 1.90 (1.40, 2.58) | <0.001 | 1.18 (0.91, 1.52) | 0.206 | 1.03 (0.76, 1.39) | 0.848 |
| 58+ years | 0.35 (0.19, 0.65) | 0.001 | 0.96 (0.68, 1.37) | 0.828 | 0.80 (0.52, 1.25) | 0.332 |
| **Marital Status** |  |  |  |  |  |  |
| Single | **1.00** |  | **1.00** |  | **1.00** |  |
| Married | 0.68 (0.56, 0.84) | <0.001 | 1.42 (1.20, 1.68) | <0.001 | 1.69 (1.36, 2.10) | <0.001 |
| Divorced/Widowed | 1.12 (0.71, 1.78) | 0.626 | 1.71 (1.15, 2.53) | 0.008 | 2.43 (1.56, 3.76) | <0.001 |
| **Working Status** |  |  |  |  |  |  |
| Working (Full time) | **1.00** |  | **1.00** |  | **1.00** |  |
| Working (Part time) | 1.23 (0.97, 1.56) | 0.089 | 0.74 (0.61, 0.89) | 0.002 | 0.88 (0.71, 1.10) | 0.281 |
| No working/Student | 0.72 (0.59, 0.89) | 0.002 | 0.40 (0.33, 0.47) | <0.001 | 0.27 (0.22, 0.34) | <0.001 |
| **Household factor** |  |  |  |  |  |  |
| **Living Arrangement** |  |  |  |  |  |  |
| Living with Family | **1.00** |  | **1.00** |  | **1.00** |  |
| Living Alone | 1.27 (1.04, 1.54) | 0.019 | 1.91 (1.61, 2.28) | <0.001 | 2.22 (1.80, 2.74) | <0.001 |
| Shared accommodation/Living with Flatmates | 1.34 (1.02, 1.77) | 0.039 | 1.77 (1.37, 2.29) | <0.001 | 1.68 (1.20, 2.35) | 0.002 |
| **Household Size** |  |  |  |  |  |  |
| 1-2 People | **1.00** |  | **1.00** |  | **1.00** |  |
| 3-4 People | 0.55 (0.45, 0.78) | <0.001 | 0.72 (0.59, 0.87) | 0.001 | 0.60 (0.48, 0.75) | <0.001 |
| **Have you been tested for COVID-19?** |  |  |  |  |  |  |
| No | **1.00** |  | **1.00** |  | **1.00** |  |
| Yes, I Tested Negative | 1.85 (1.38, 2.49) | <0.001 | 2.52 (1.94, 3.26) | <0.001 | 2.24 (1.67, 3.02) | <0.001 |
| Yes. I Tested Positive | 1.48 (0.87, 2.54) | 0.152 | 2.08 (1.31, 3.28) | 0.002 | 1.92 (1.17, 3.16) | 0.010 |
| **Compliance with Public health measures** |  |  |  |  |  |  |
| **Are You Currently in Self-Quarantine Since Past Seven Days?** |  |  |  |  |  |  |
| No | **1.00** |  | **1.00** |  | **1.00** |  |
| Yes | 0.77 (0.66, 0.91) | 0.002 | 1.41 (1.21, 1.64) | <0.001 | 1.89 (1.53, 2.33) | <0.001 |
| **Avoid Public Transport** |  |  |  |  |  |  |
| Yes | ***1.00*** |  | ***1.00*** |  | ***1.00*** |  |
| No | 0.91 (0.62, 1.33) | 0.626 | 2.57 (1.86, 3.56) | <0.001 | 2.52 (1.70, 3.73) | <0.001 |
| **Avoid Shaking Hands** |  |  |  |  |  |  |
| Yes | ***1.00*** |  | ***1.00*** |  | ***1.00*** |  |
| No | 0.69 (0.48, 1.00) | 0.051 | 1.08 (0.78, 1.48) | 0.651 | 1.76 (1.20, 2.56) | 0.003 |
| **Wearing Face Mask when going out** |  |  |  |  |  |  |
| Yes | ***1.00*** |  | ***1.00*** |  | ***1.00*** |  |
| No | 0.92 (0.66, 1.28) | 0.634 | 1.28 (0.97, 1.71) | 0.081 | 1.39 (0.99, 1.96) | 0.057 |
| **Avoid Large Gatherings** |  |  |  |  |  |  |
| Yes | ***1.00*** |  | ***1.00*** |  | ***1.00*** |  |
| No | 1.53 (1.10, 2.12) | 0.011 | 0.62 (0.46, 0.82) | 0.001 | 0.67 (0.48, 0.95) | 0.023 |
| **Advocating People About the Health Risk Related to COVID-19** |  |  |  |  |  |  |
| Yes | ***1.00*** |  | ***1.00*** |  | ***1.00*** |  |
| No | 0.76 (0.61, 0.95) | 0.017 | 0.50 (0.41, 0.62) | <0.001 | 0.56 (0.43, 0.73) | <0.001 |
| **Health Condition experienced in the Last Week** |  |  |  |  |  |  |
| **Fever** |  |  |  |  |  |  |
| No | ***1.00*** |  | ***1.00*** |  | ***1.00*** |  |
| Yes | 1.25 (0.99, 1.57) | 0.051 | 0.90 (0.74, 1.10) | 0.290 | 1.37 (1.09, 1.72) | 0.006 |
| **Chills** |  |  |  |  |  |  |
| No | ***1.00*** |  | ***1.00*** |  | ***1.00*** |  |
| Yes | 1.32 (1.01, 1.72) | 0.041 | 2.05 (1.65, 2.56) | <0.001 | 2.19 (1.72, 2.78) | <0.001 |
| **Headache** |  |  |  |  |  |  |
| No | ***1.00*** |  | ***1.00*** |  | ***1.00*** |  |
| Yes | 1.15 (0.98, 1.36) | 0.089 | 1.43 (1.25, 1.64) | <0.001 | 1.13 (0.96, 1.35) | 0.144 |
| **Cough** |  |  |  |  |  |  |
| No | **1.00** |  | **1.00** |  | **1.00** |  |
| Yes | 2.55 (2.14, 3.03) | <0.001 | 2.45 (2.10, 2.86) | <0.001 | 1.68 (1.39, 2.02) | <0.001 |
| **Breathing Difficulty** |  |  |  |  |  |  |
| No | **1.00** |  | **1.00** |  | **1.00** |  |
| Yes | 3.29 (2.48, 4.35) | <0.001 | 3.15 (2.47, 4.03) | <0.001 | 4.43 (3.41, 5.76) | <0.001 |
| **Dizziness** |  |  |  |  |  |  |
| No | **1.00** |  | **1.00** |  | **1.00** |  |
| Yes | 1.92 (1.53, 2.40) | <0.001 | 2.42 (1.99, 2.93) | <0.001 | 1.86 (1.49, 2.32) | 0.001 |
| **Sore Throat** |  |  |  |  |  |  |
| No | **1.00** |  | **1.00** |  | **1.00** |  |
| Yes | 1.84 (1.48, 2.27) | <0.001 | 2.26 (1.89, 2.70) | <0.001 | 2.15 (1.75, 2.65) | <0.001 |
| Adjusted odd ratios (aOR); 95% confidence intervals (CI) |  |  |  |  |  |  |

**Figure 2.**
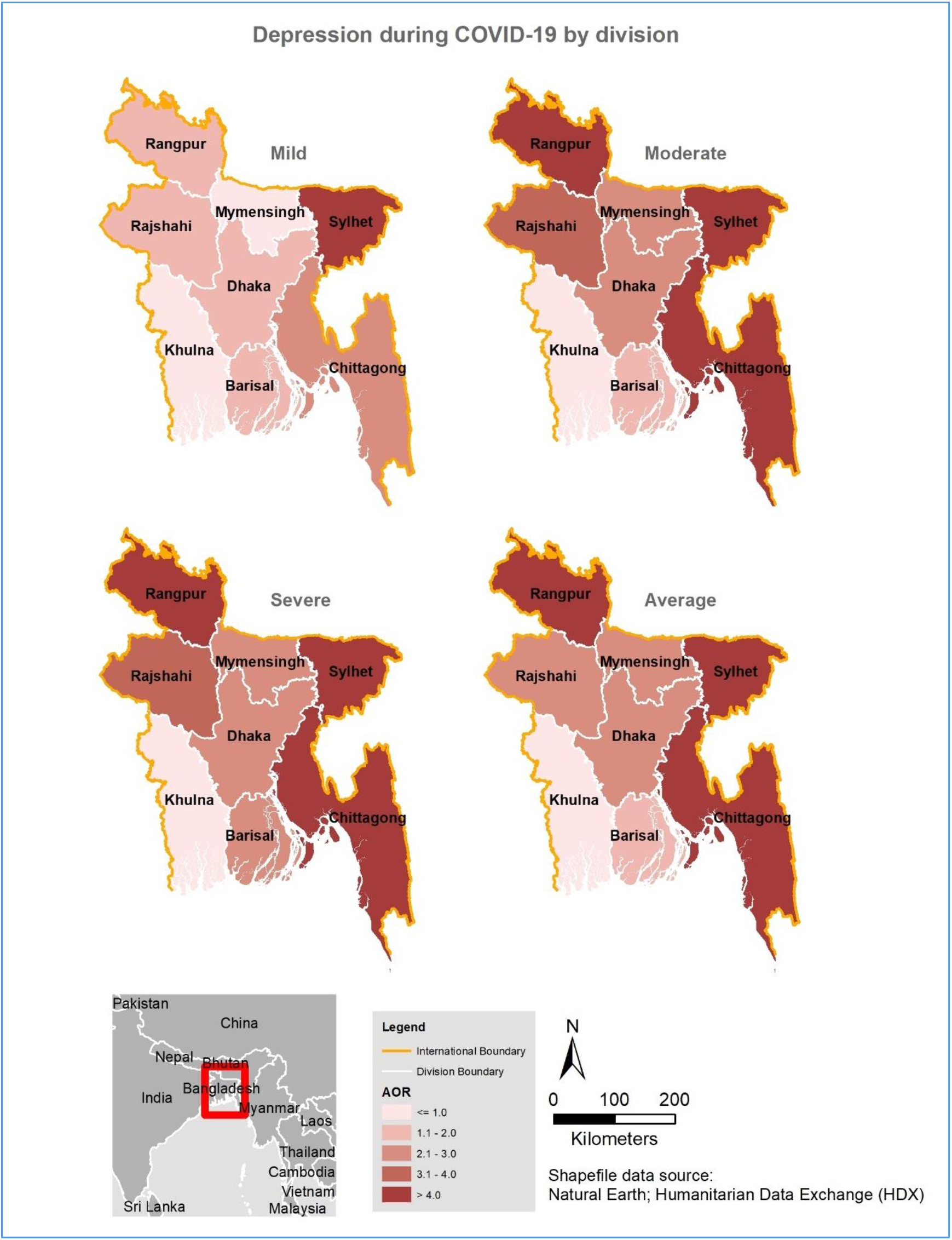
Spatial distribution of depression during COVID-19 in Bangladesh by division

Table 3 presents the multivariate analysis of factors associated with anxiety among the participants. Their distribution by region is shown in Figure 3, revealing that the odds for feeling severely anxious increased by more than 3 folds among respondents from four of the eight regions (see Figure 3 for details). From the Table, females, those who were divorced or widowed, those who lived in shared accommodation during the lockdown, individuals who experienced self-quarantine, and those who experienced fever and cough a couple of weeks before data collection, were more likely to experience mild-severe symptoms of anxiety, while older participants experienced a lower risk of depression at all levels. Compared to those from Khulna division, participants who lived in other divisions particularly Chittagong [adjusted OR, 6.26, 95%CI, 4.40-8.90], Mymensing [adjusted OR, 5.26, 95%CI, 3.04-9.12] and Rangpur [adjusted OR, 5.86, 95%CI, 3.64-9.42] had a higher risk of severe symptoms of depression (see Figure 4) as well as those who lived alone [adjusted OR, 1.70, 95%CI, 1.35-2.16] during the lockdown. Those with confirmed or suspected cases of COVID-19 were more about two times more likely to experience severe symptoms of depression compared to those who were not tested. Other symptoms of COVID-19 were also significantly associated with some degree of depression in this study.

**Table 3.**
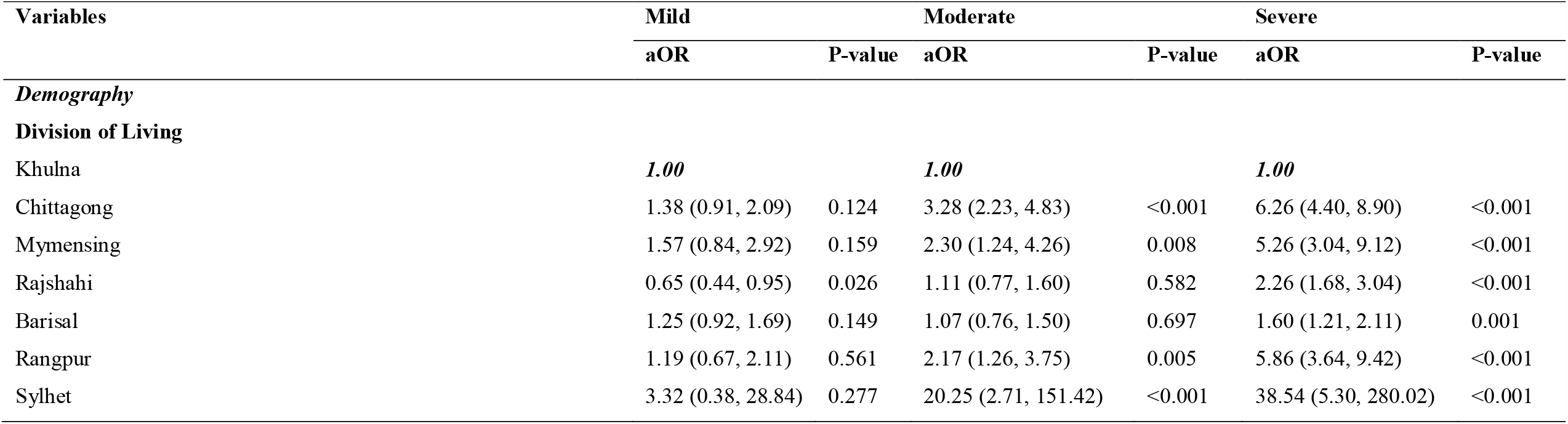
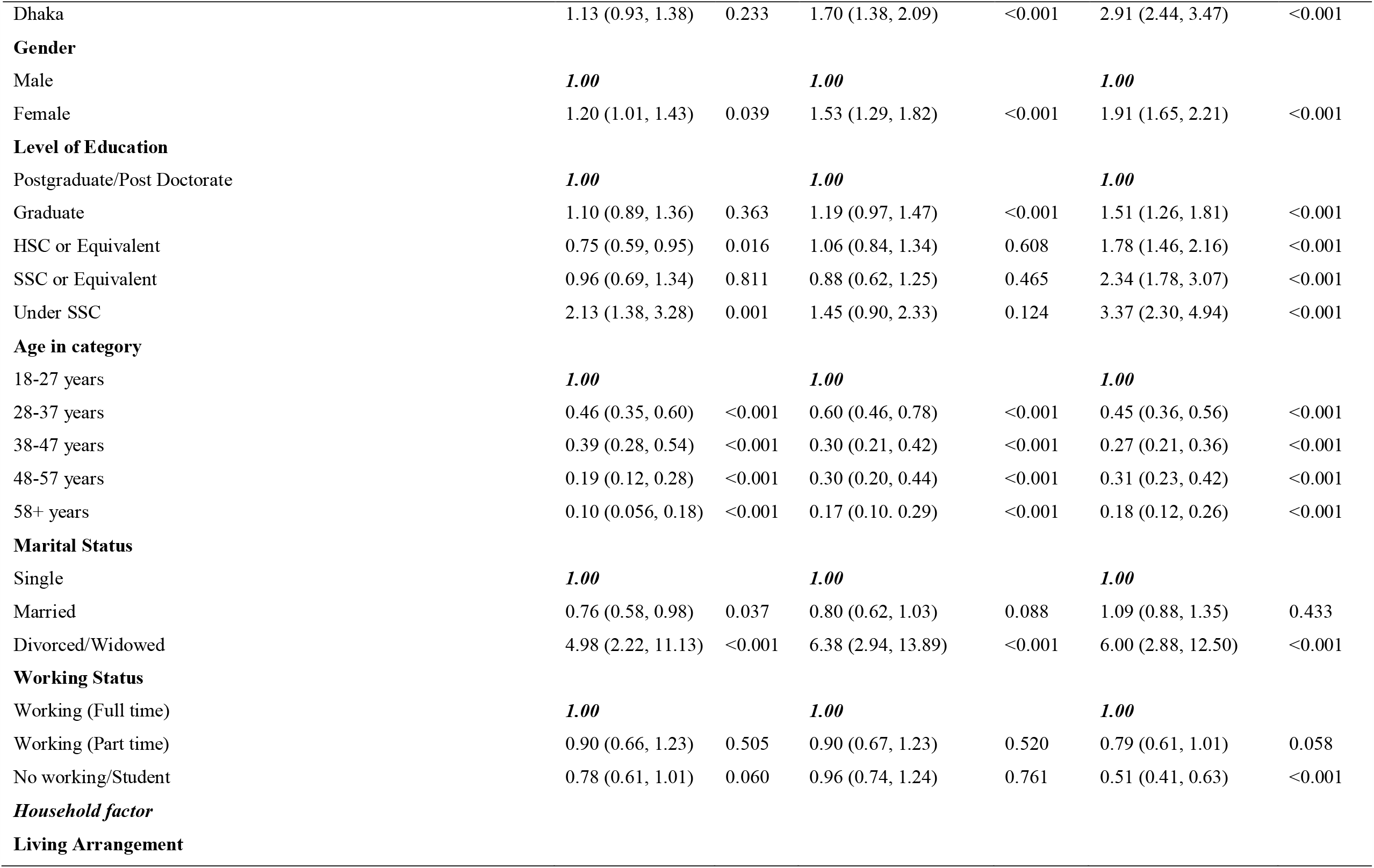
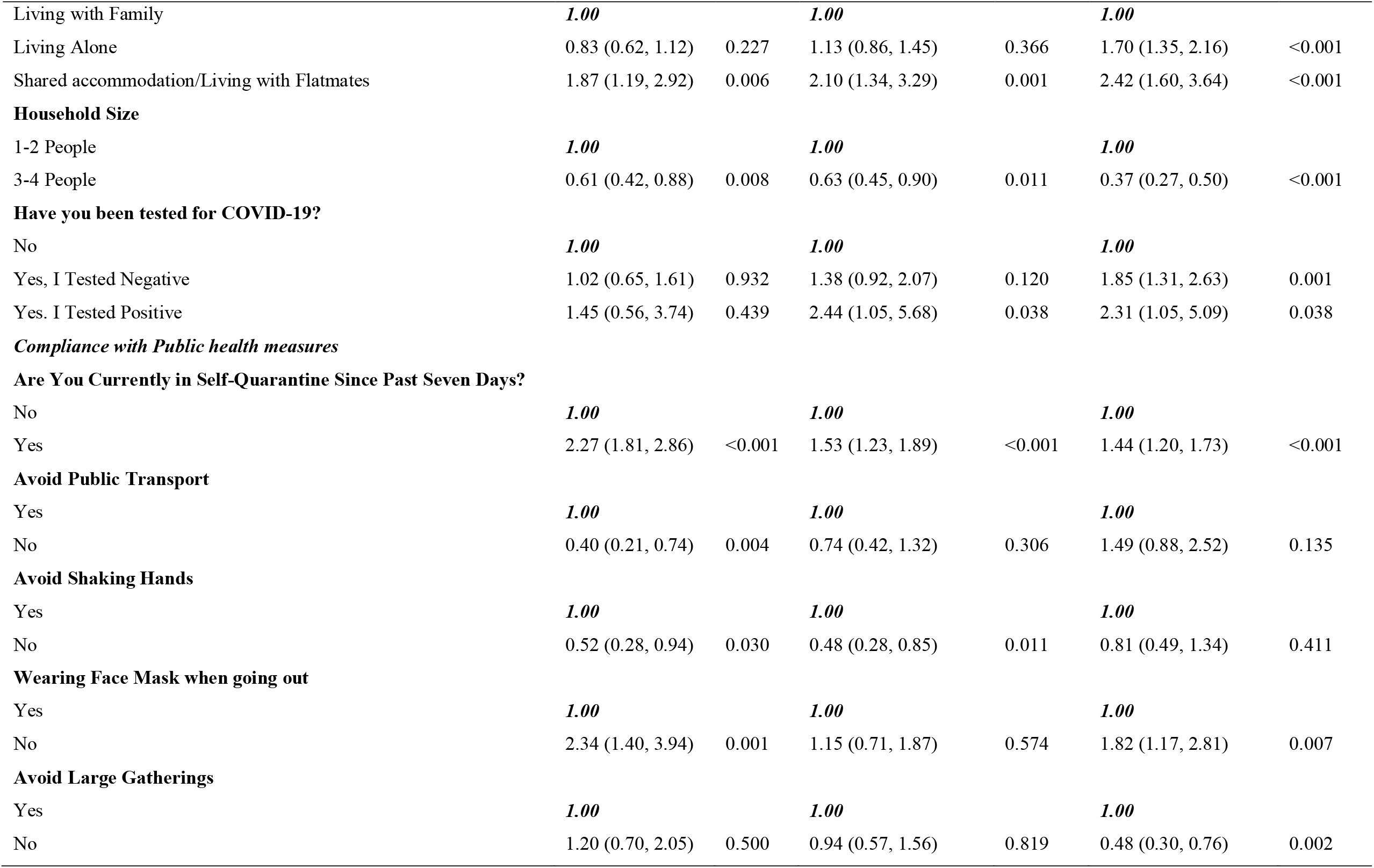
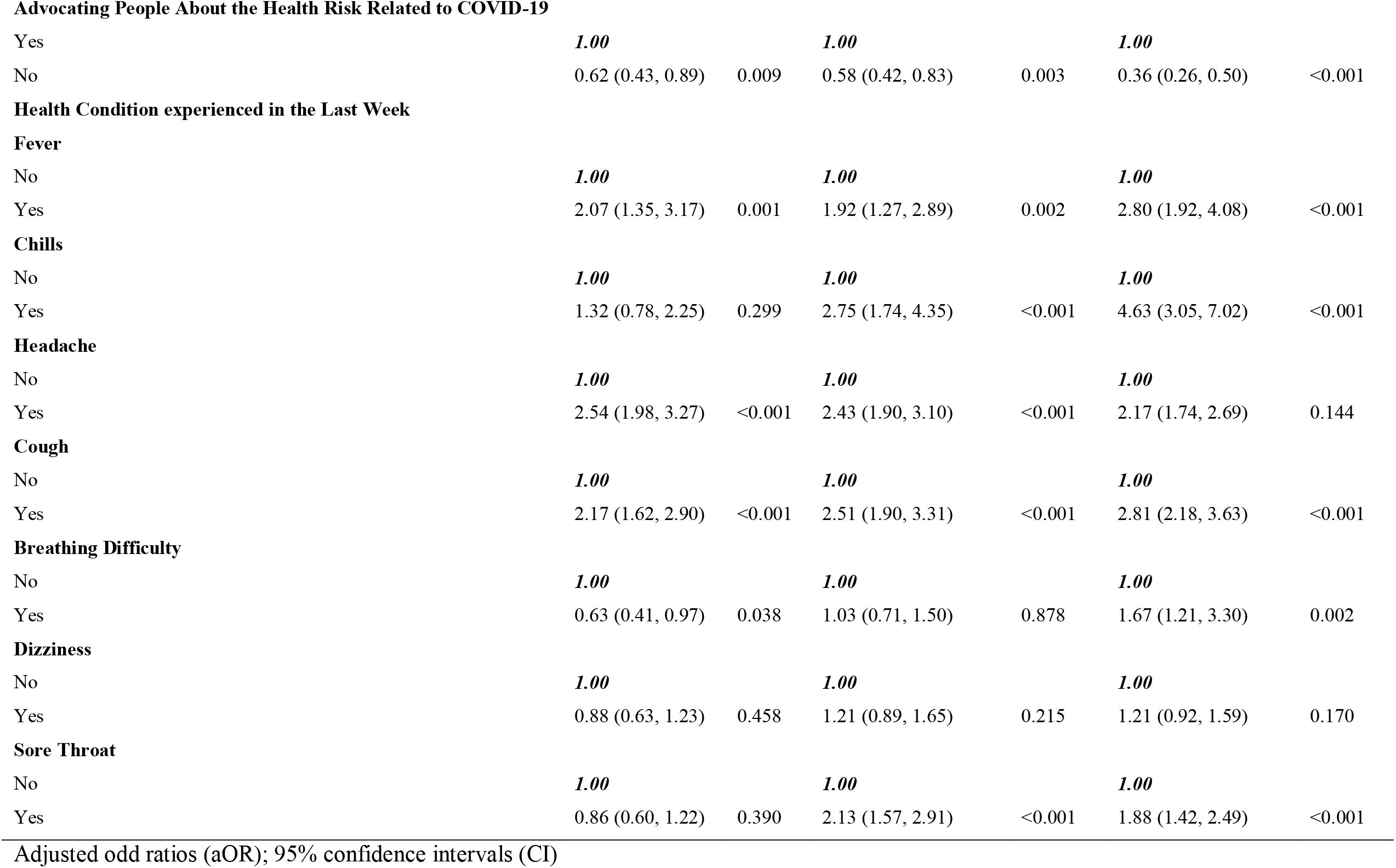
Factors associated with mild, moderate, and severe anxiety during COVID-19 in Bangladesh.

| Variables | Mild |  | Moderate |  | Severe |  |
| --- | --- | --- | --- | --- | --- | --- |
|  | aOR | P-value | aOR | P-value | aOR | P-value |
| **Demography** |  |  |  |  |  |  |
| **Division of Living** |  |  |  |  |  |  |
| Khulna | **1.00** |  | **1.00** |  | **1.00** |  |
| Chittagong | 1.38 (0.91, 2.09) | 0.124 | 3.28 (2.23, 4.83) | <0.001 | 6.26 (4.40, 8.90) | <0.001 |
| Mymensing | 1.57 (0.84, 2.92) | 0.159 | 2.30 (1.24, 4.26) | 0.008 | 5.26 (3.04, 9.12) | <0.001 |
| Rajshahi | 0.65 (0.44, 0.95) | 0.026 | 1.11 (0.77, 1.60) | 0.582 | 2.26 (1.68, 3.04) | <0.001 |
| Barisal | 1.25 (0.92, 1.69) | 0.149 | 1.07 (0.76, 1.50) | 0.697 | 1.60 (1.21, 2.11) | 0.001 |
| Rangpur | 1.19 (0.67, 2.11) | 0.561 | 2.17 (1.26, 3.75) | 0.005 | 5.86 (3.64, 9.42) | <0.001 |
| Sylhet | 3.32 (0.38, 28.84) | 0.277 | 20.25 (2.71, 151.42) | <0.001 | 38.54 (5.30, 280.02) | <0.001 |
| Dhaka | 1.13 (0.93, 1.38) | 0.233 | 1.70 (1.38, 2.09) | <0.001 | 2.91 (2.44, 3.47) | <0.001 |
| **Gender** |  |  |  |  |  |  |
| Male | *1.00* |  | *1.00* |  | *1.00* |  |
| Female | 1.20 (1.01, 1.43) | 0.039 | 1.53 (1.29, 1.82) | <0.001 | 1.91 (1.65, 2.21) | <0.001 |
| **Level of Education** |  |  |  |  |  |  |
| Postgraduate/Post Doctorate | *1.00* |  | *1.00* |  | *1.00* |  |
| Graduate | 1.10 (0.89, 1.36) | 0.363 | 1.19 (0.97, 1.47) | <0.001 | 1.51 (1.26, 1.81) | <0.001 |
| HSC or Equivalent | 0.75 (0.59, 0.95) | 0.016 | 1.06 (0.84, 1.34) | 0.608 | 1.78 (1.46, 2.16) | <0.001 |
| SSC or Equivalent | 0.96 (0.69, 1.34) | 0.811 | 0.88 (0.62, 1.25) | 0.465 | 2.34 (1.78, 3.07) | <0.001 |
| Under SSC | 2.13 (1.38, 3.28) | 0.001 | 1.45 (0.90, 2.33) | 0.124 | 3.37 (2.30, 4.94) | <0.001 |
| **Age in category** |  |  |  |  |  |  |
| 18-27 years | *1.00* |  | *1.00* |  | *1.00* |  |
| 28-37 years | 0.46 (0.35, 0.60) | <0.001 | 0.60 (0.46, 0.78) | <0.001 | 0.45 (0.36, 0.56) | <0.001 |
| 38-47 years | 0.39 (0.28, 0.54) | <0.001 | 0.30 (0.21, 0.42) | <0.001 | 0.27 (0.21, 0.36) | <0.001 |
| 48-57 years | 0.19 (0.12, 0.28) | <0.001 | 0.30 (0.20, 0.44) | <0.001 | 0.31 (0.23, 0.42) | <0.001 |
| 58+ years | 0.10 (0.056, 0.18) | <0.001 | 0.17 (0.10, 0.29) | <0.001 | 0.18 (0.12, 0.26) | <0.001 |
| **Marital Status** |  |  |  |  |  |  |
| Single | *1.00* |  | *1.00* |  | *1.00* |  |
| Married | 0.76 (0.58, 0.98) | 0.037 | 0.80 (0.62, 1.03) | 0.088 | 1.09 (0.88, 1.35) | 0.433 |
| Divorced/Widowed | 4.98 (2.22, 11.13) | <0.001 | 6.38 (2.94, 13.89) | <0.001 | 6.00 (2.88, 12.50) | <0.001 |
| **Working Status** |  |  |  |  |  |  |
| Working (Full time) | *1.00* |  | *1.00* |  | *1.00* |  |
| Working (Part time) | 0.90 (0.66, 1.23) | 0.505 | 0.90 (0.67, 1.23) | 0.520 | 0.79 (0.61, 1.01) | 0.058 |
| No working/Student | 0.78 (0.61, 1.01) | 0.060 | 0.96 (0.74, 1.24) | 0.761 | 0.51 (0.41, 0.63) | <0.001 |
| **Household factor** |  |  |  |  |  |  |
| **Living Arrangement** |  |  |  |  |  |  |
| Living with Family | **1.00** |  | **1.00** |  | **1.00** |  |
| Living Alone | 0.83 (0.62, 1.12) | 0.227 | 1.13 (0.86, 1.45) | 0.366 | 1.70 (1.35, 2.16) | <0.001 |
| Shared accommodation/Living with Flatmates | 1.87 (1.19, 2.92) | 0.006 | 2.10 (1.34, 3.29) | 0.001 | 2.42 (1.60, 3.64) | <0.001 |
| **Household Size** |  |  |  |  |  |  |
| 1-2 People | **1.00** |  | **1.00** |  | **1.00** |  |
| 3-4 People | 0.61 (0.42, 0.88) | 0.008 | 0.63 (0.45, 0.90) | 0.011 | 0.37 (0.27, 0.50) | <0.001 |
| **Have you been tested for COVID-19?** |  |  |  |  |  |  |
| No | **1.00** |  | **1.00** |  | **1.00** |  |
| Yes, I Tested Negative | 1.02 (0.65, 1.61) | 0.932 | 1.38 (0.92, 2.07) | 0.120 | 1.85 (1.31, 2.63) | 0.001 |
| Yes, I Tested Positive | 1.45 (0.56, 3.74) | 0.439 | 2.44 (1.05, 5.68) | 0.038 | 2.31 (1.05, 5.09) | 0.038 |
| **Compliance with Public health measures** |  |  |  |  |  |  |
| **Are You Currently in Self-Quarantine Since Past Seven Days?** |  |  |  |  |  |  |
| No | **1.00** |  | **1.00** |  | **1.00** |  |
| Yes | 2.27 (1.81, 2.86) | <0.001 | 1.53 (1.23, 1.89) | <0.001 | 1.44 (1.20, 1.73) | <0.001 |
| **Avoid Public Transport** |  |  |  |  |  |  |
| Yes | **1.00** |  | **1.00** |  | **1.00** |  |
| No | 0.40 (0.21, 0.74) | 0.004 | 0.74 (0.42, 1.32) | 0.306 | 1.49 (0.88, 2.52) | 0.135 |
| **Avoid Shaking Hands** |  |  |  |  |  |  |
| Yes | **1.00** |  | **1.00** |  | **1.00** |  |
| No | 0.52 (0.28, 0.94) | 0.030 | 0.48 (0.28, 0.85) | 0.011 | 0.81 (0.49, 1.34) | 0.411 |
| **Wearing Face Mask when going out** |  |  |  |  |  |  |
| Yes | **1.00** |  | **1.00** |  | **1.00** |  |
| No | 2.34 (1.40, 3.94) | 0.001 | 1.15 (0.71, 1.87) | 0.574 | 1.82 (1.17, 2.81) | 0.007 |
| **Avoid Large Gatherings** |  |  |  |  |  |  |
| Yes | **1.00** |  | **1.00** |  | **1.00** |  |
| No | 1.20 (0.70, 2.05) | 0.500 | 0.94 (0.57, 1.56) | 0.819 | 0.48 (0.30, 0.76) | 0.002 |
| **Advocating People About the Health Risk Related to COVID-19** |  |  |  |  |  |  |
| Yes | **1.00** |  | **1.00** |  | **1.00** |  |
| No | 0.62 (0.43, 0.89) | 0.009 | 0.58 (0.42, 0.83) | 0.003 | 0.36 (0.26, 0.50) | <0.001 |
| **Health Condition experienced in the Last Week** |  |  |  |  |  |  |
| **Fever** |  |  |  |  |  |  |
| No | **1.00** |  | **1.00** |  | **1.00** |  |
| Yes | 2.07 (1.35, 3.17) | 0.001 | 1.92 (1.27, 2.89) | 0.002 | 2.80 (1.92, 4.08) | <0.001 |
| **Chills** |  |  |  |  |  |  |
| No | **1.00** |  | **1.00** |  | **1.00** |  |
| Yes | 1.32 (0.78, 2.25) | 0.299 | 2.75 (1.74, 4.35) | <0.001 | 4.63 (3.05, 7.02) | <0.001 |
| **Headache** |  |  |  |  |  |  |
| No | **1.00** |  | **1.00** |  | **1.00** |  |
| Yes | 2.54 (1.98, 3.27) | <0.001 | 2.43 (1.90, 3.10) | <0.001 | 2.17 (1.74, 2.69) | 0.144 |
| **Cough** |  |  |  |  |  |  |
| No | **1.00** |  | **1.00** |  | **1.00** |  |
| Yes | 2.17 (1.62, 2.90) | <0.001 | 2.51 (1.90, 3.31) | <0.001 | 2.81 (2.18, 3.63) | <0.001 |
| **Breathing Difficulty** |  |  |  |  |  |  |
| No | **1.00** |  | **1.00** |  | **1.00** |  |
| Yes | 0.63 (0.41, 0.97) | 0.038 | 1.03 (0.71, 1.50) | 0.878 | 1.67 (1.21, 3.30) | 0.002 |
| **Dizziness** |  |  |  |  |  |  |
| No | **1.00** |  | **1.00** |  | **1.00** |  |
| Yes | 0.88 (0.63, 1.23) | 0.458 | 1.21 (0.89, 1.65) | 0.215 | 1.21 (0.92, 1.59) | 0.170 |
| **Sore Throat** |  |  |  |  |  |  |
| No | **1.00** |  | **1.00** |  | **1.00** |  |
| Yes | 0.86 (0.60, 1.22) | 0.390 | 2.13 (1.57, 2.91) | <0.001 | 1.88 (1.42, 2.49) | <0.001 |

Adjusted odd ratios (aOR); 95% confidence intervals (CI)

**Figure 3.**
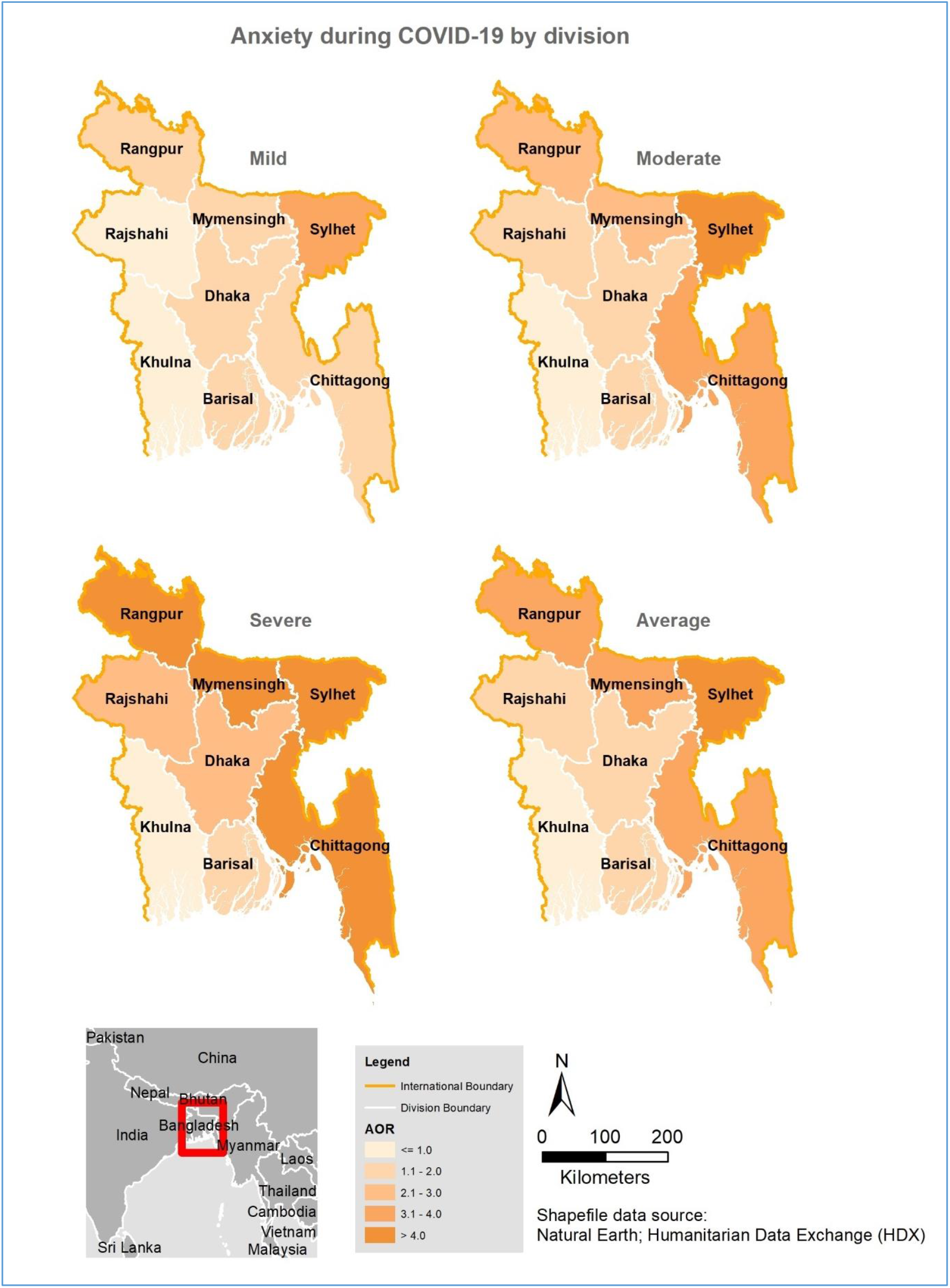
Spatial distribution of anxiety during COVID-19 in Bangladesh by division

**Figure 4.**
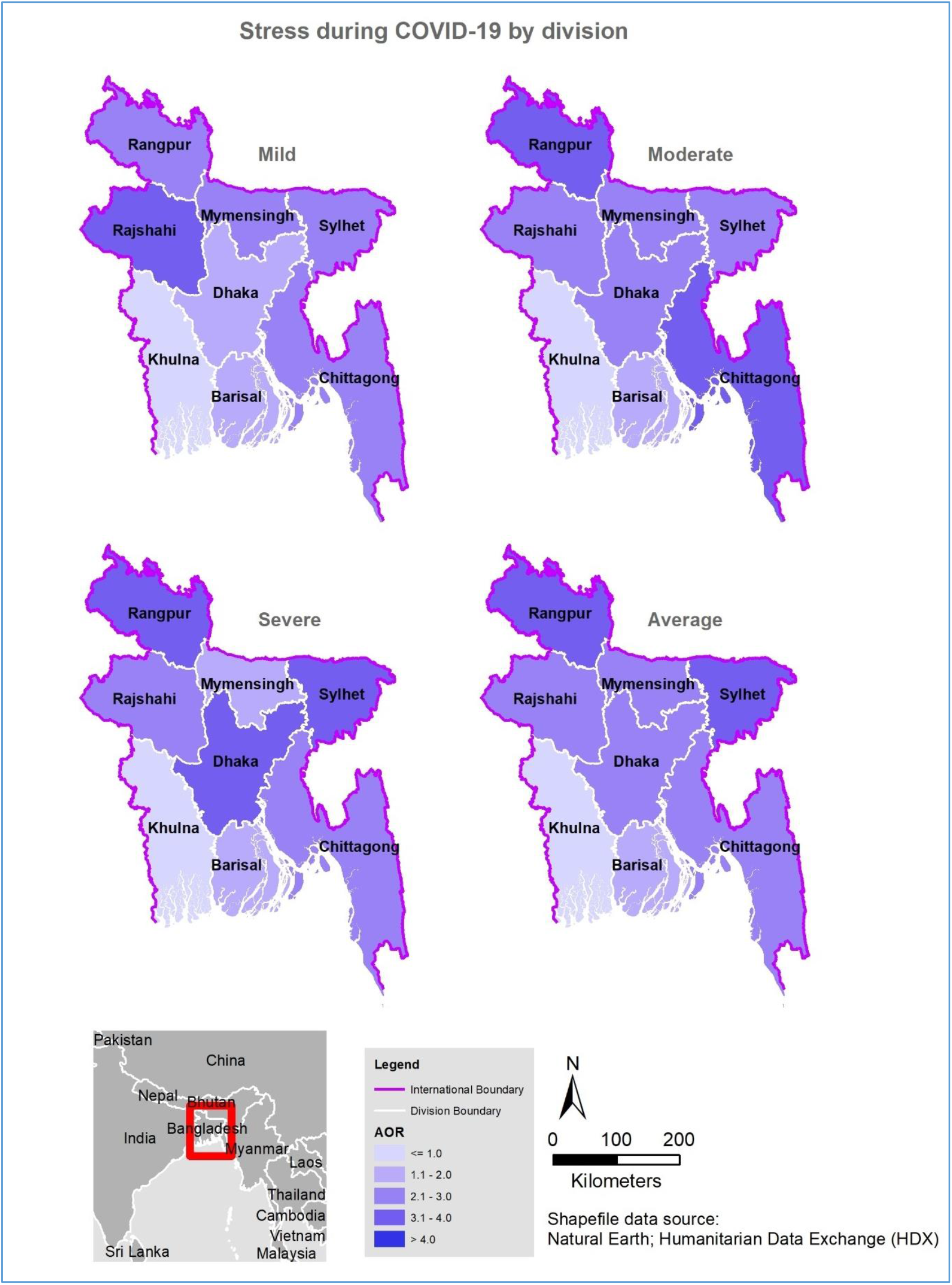
Spatial distribution of stress during COVID-19 in Bangladesh by division

The factors associated with stress in the multivariate analysis are shown in Table 4. Females, those living outside of Khulna division, participants with lower than postgraduate education, older people (>27years), participants who were married, those who lived alone, and participants who had experienced self-quarantine and any of the health symptoms in this study (except for fever) were more likely to experience symptoms of stress at all levels. Participants who were divorced/widowed [adjusted OR, 1.54, 95%CI, 1.06-2.24], those who lived in shared accommodations [adjusted OR, 2.26, 95%CI, 1.69-3.02] as well as individuals who failed to comply with the precautionary measures advising people to avoid traveling by public transport and/or shake hands [adjusted OR, 1.63, 95%CI, 1.15-2.30] had a remarkably higher risk of moderate and severe stress symptoms [adjusted OR, 4.02, 95%CI, 2.74-5.91 and 1.63, 95%CI, 1.15-2.30, respectively for severe stress], while severe stress symptoms were also found among those who failed to wear face masks when going out [adjusted OR, 1.64, 95%CI, 1.19-2.25].

**Table 4:**
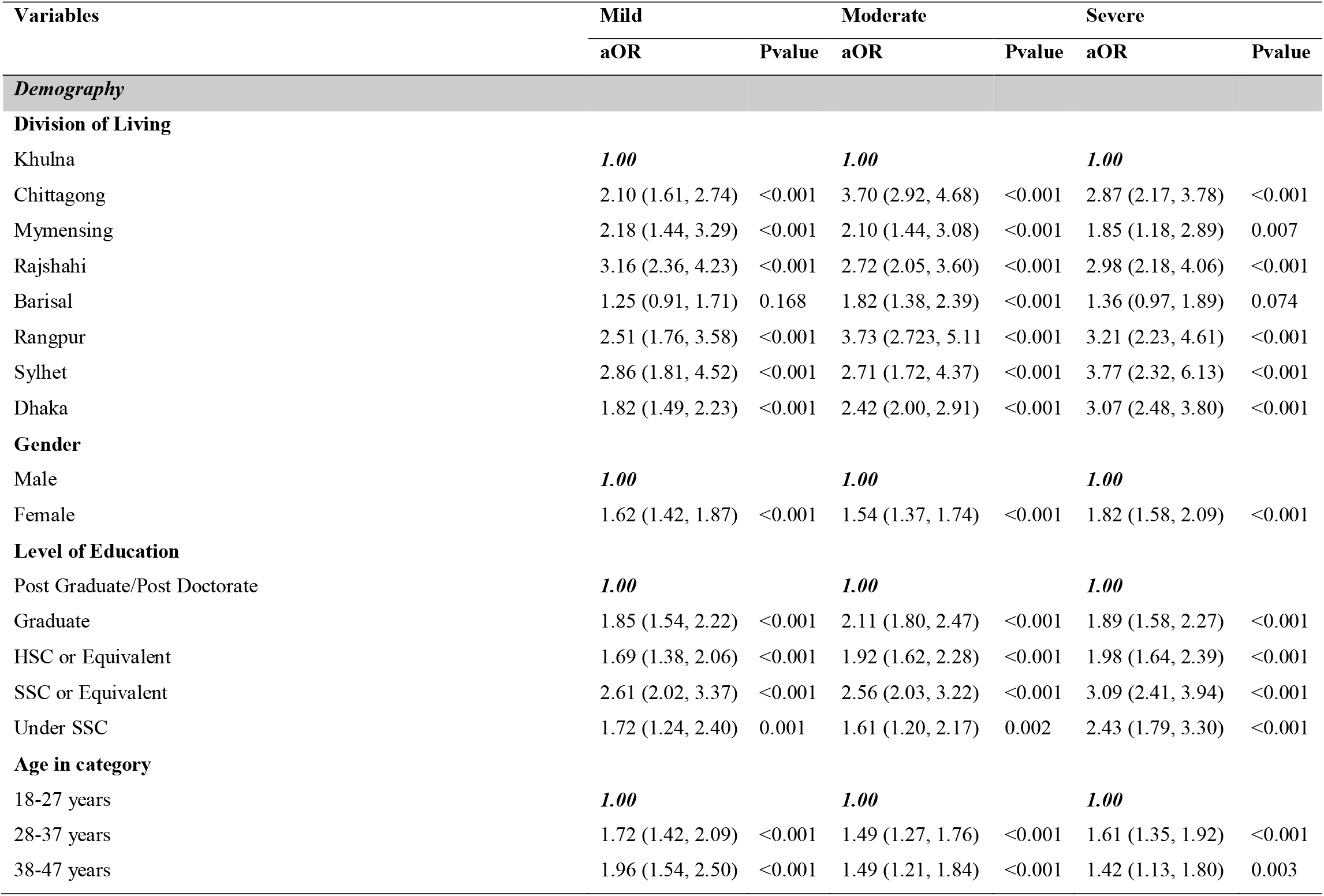
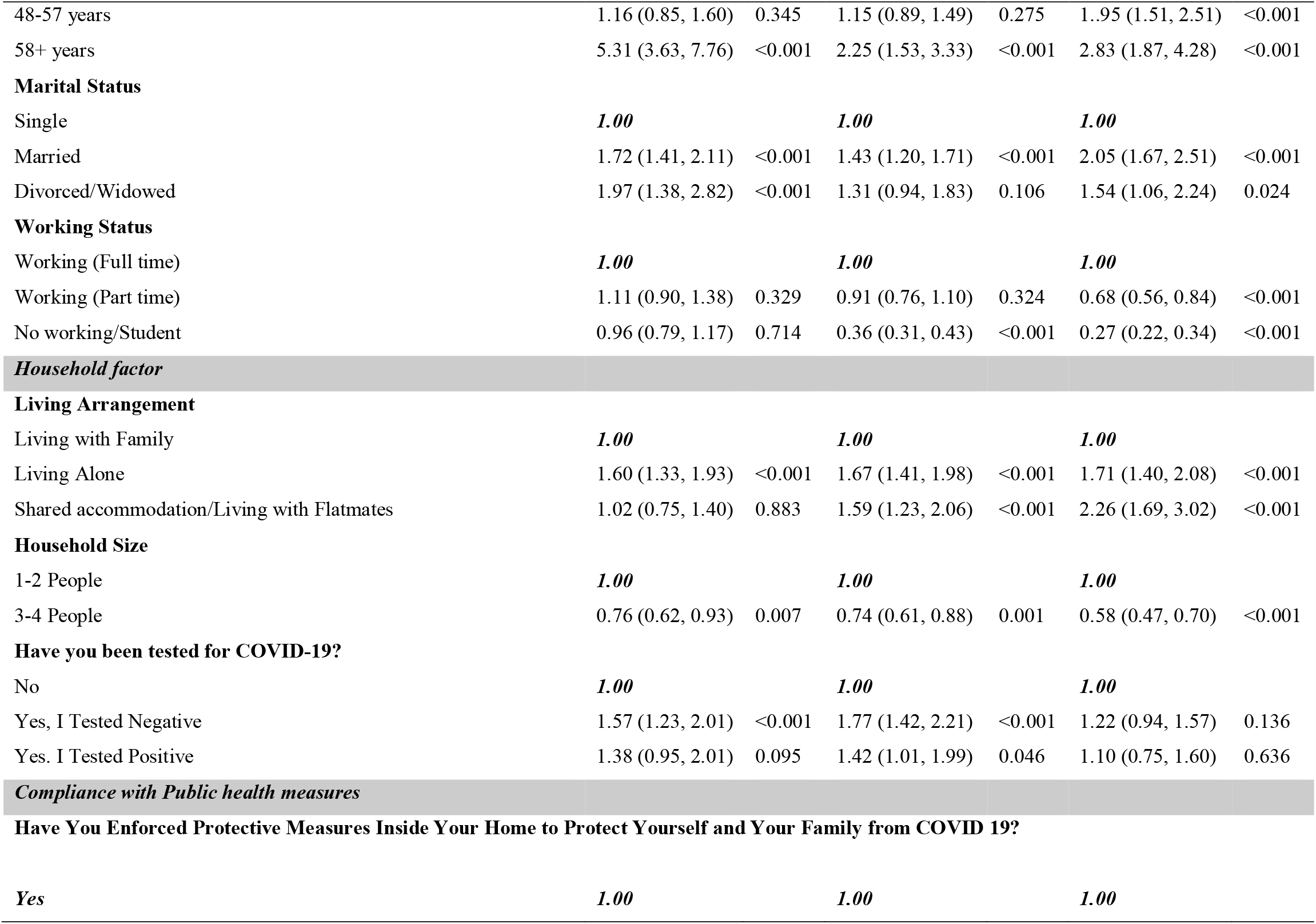
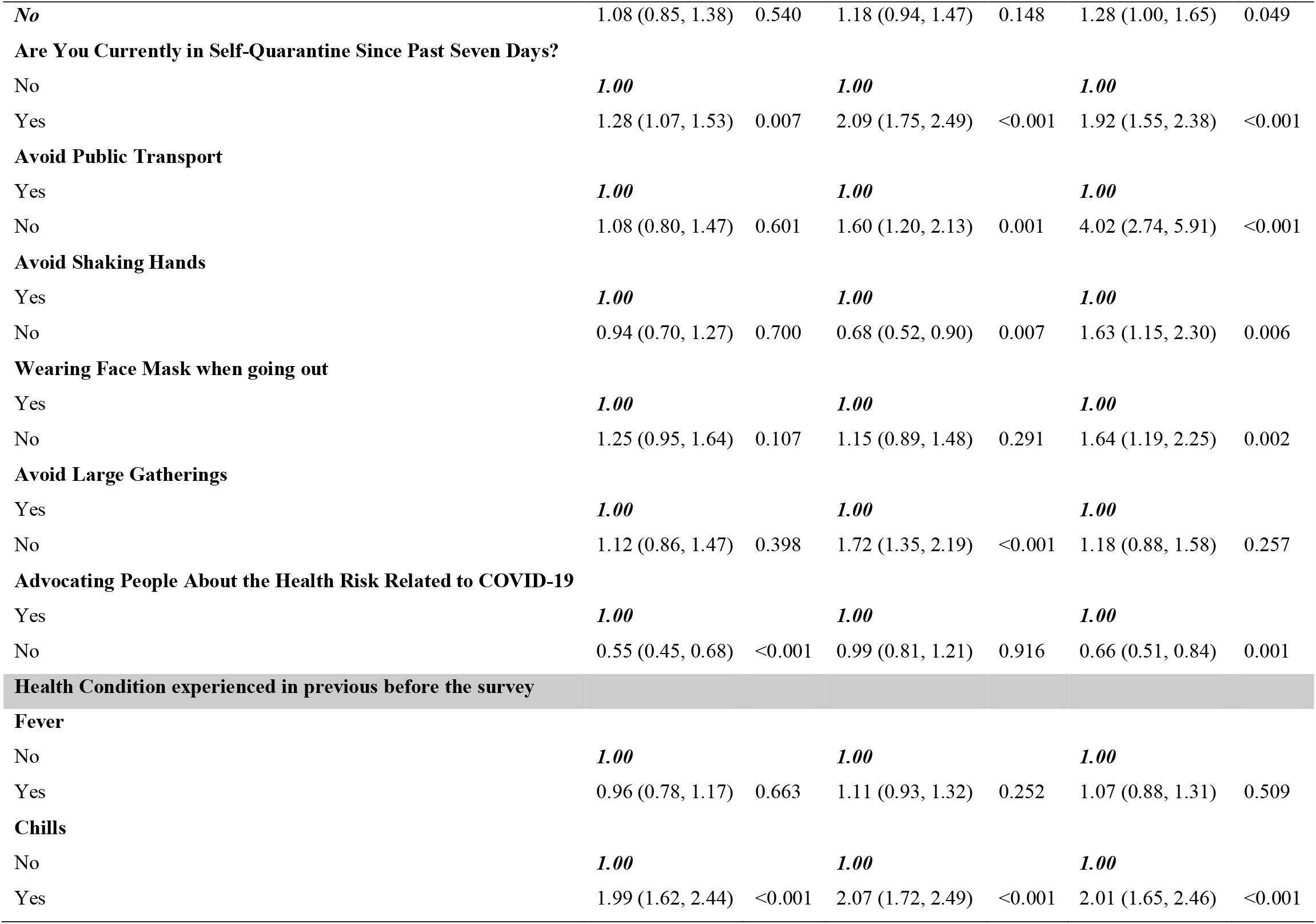
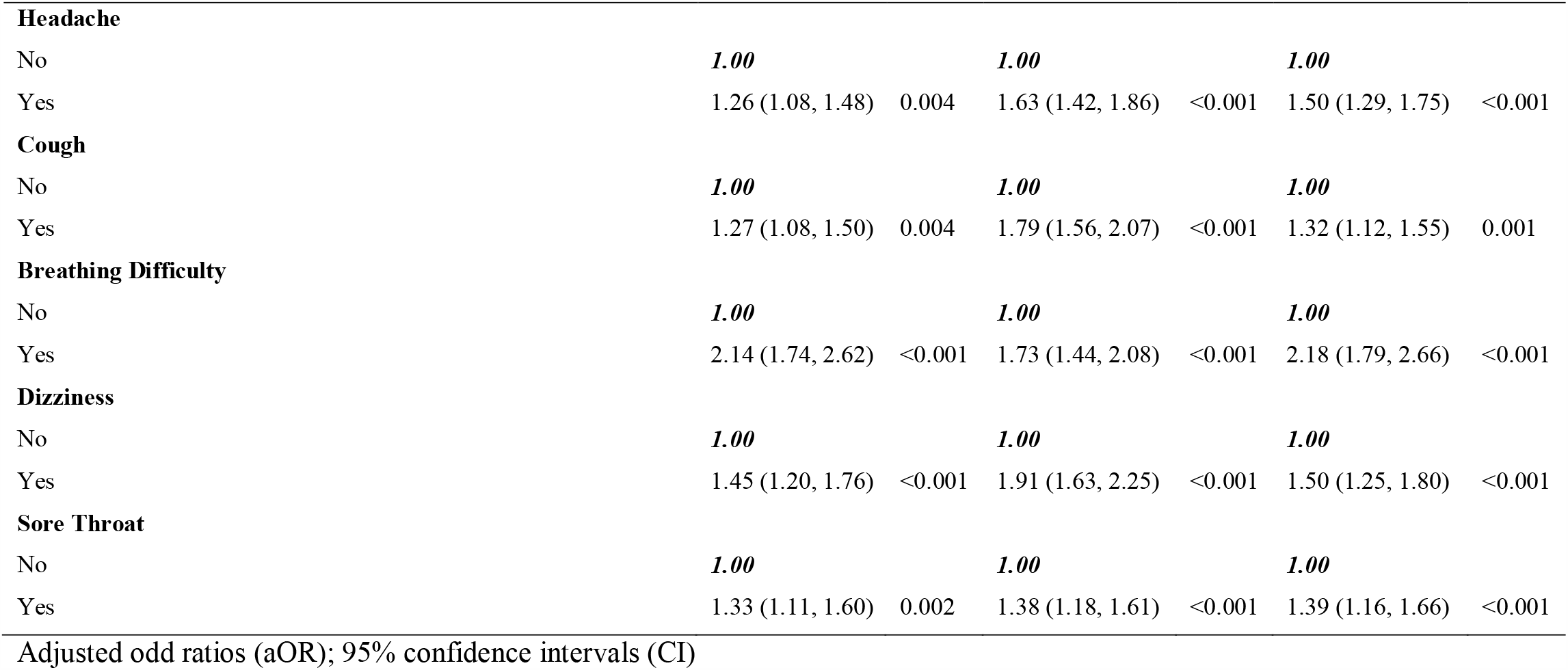
Factors associated with mild, moderate, and severe Stress during COVID-19 in Bangladesh.

| Variables | Mild |  | Moderate |  | Severe |  |
| --- | --- | --- | --- | --- | --- | --- |
|  | aOR | Pvalue | aOR | Pvalue | aOR | Pvalue |
| **Demography** |  |  |  |  |  |  |
| **Division of Living** |  |  |  |  |  |  |
| Khulna | **1.00** |  | **1.00** |  | **1.00** |  |
| Chittagong | 2.10 (1.61, 2.74) | <0.001 | 3.70 (2.92, 4.68) | <0.001 | 2.87 (2.17, 3.78) | <0.001 |
| Mymensing | 2.18 (1.44, 3.29) | <0.001 | 2.10 (1.44, 3.08) | <0.001 | 1.85 (1.18, 2.89) | 0.007 |
| Rajshahi | 3.16 (2.36, 4.23) | <0.001 | 2.72 (2.05, 3.60) | <0.001 | 2.98 (2.18, 4.06) | <0.001 |
| Barisal | 1.25 (0.91, 1.71) | 0.168 | 1.82 (1.38, 2.39) | <0.001 | 1.36 (0.97, 1.89) | 0.074 |
| Rangpur | 2.51 (1.76, 3.58) | <0.001 | 3.73 (2.723, 5.11) | <0.001 | 3.21 (2.23, 4.61) | <0.001 |
| Sylhet | 2.86 (1.81, 4.52) | <0.001 | 2.71 (1.72, 4.37) | <0.001 | 3.77 (2.32, 6.13) | <0.001 |
| Dhaka | 1.82 (1.49, 2.23) | <0.001 | 2.42 (2.00, 2.91) | <0.001 | 3.07 (2.48, 3.80) | <0.001 |
| **Gender** |  |  |  |  |  |  |
| Male | **1.00** |  | **1.00** |  | **1.00** |  |
| Female | 1.62 (1.42, 1.87) | <0.001 | 1.54 (1.37, 1.74) | <0.001 | 1.82 (1.58, 2.09) | <0.001 |
| **Level of Education** |  |  |  |  |  |  |
| Post Graduate/Post Doctorate | **1.00** |  | **1.00** |  | **1.00** |  |
| Graduate | 1.85 (1.54, 2.22) | <0.001 | 2.11 (1.80, 2.47) | <0.001 | 1.89 (1.58, 2.27) | <0.001 |
| HSC or Equivalent | 1.69 (1.38, 2.06) | <0.001 | 1.92 (1.62, 2.28) | <0.001 | 1.98 (1.64, 2.39) | <0.001 |
| SSC or Equivalent | 2.61 (2.02, 3.37) | <0.001 | 2.56 (2.03, 3.22) | <0.001 | 3.09 (2.41, 3.94) | <0.001 |
| Under SSC | 1.72 (1.24, 2.40) | 0.001 | 1.61 (1.20, 2.17) | 0.002 | 2.43 (1.79, 3.30) | <0.001 |
| **Age in category** |  |  |  |  |  |  |
| 18-27 years | **1.00** |  | **1.00** |  | **1.00** |  |
| 28-37 years | 1.72 (1.42, 2.09) | <0.001 | 1.49 (1.27, 1.76) | <0.001 | 1.61 (1.35, 1.92) | <0.001 |
| 38-47 years | 1.96 (1.54, 2.50) | <0.001 | 1.49 (1.21, 1.84) | <0.001 | 1.42 (1.13, 1.80) | 0.003 |
| 48-57 years | 1.16 (0.85, 1.60) | 0.345 | 1.15 (0.89, 1.49) | 0.275 | 1..95 (1.51, 2.51) | <0.001 |
| 58+ years | 5.31 (3.63, 7.76) | <0.001 | 2.25 (1.53, 3.33) | <0.001 | 2.83 (1.87, 4.28) | <0.001 |
| **Marital Status** |  |  |  |  |  |  |
| Single | **1.00** |  | **1.00** |  | **1.00** |  |
| Married | 1.72 (1.41, 2.11) | <0.001 | 1.43 (1.20, 1.71) | <0.001 | 2.05 (1.67, 2.51) | <0.001 |
| Divorced/Widowed | 1.97 (1.38, 2.82) | <0.001 | 1.31 (0.94, 1.83) | 0.106 | 1.54 (1.06, 2.24) | 0.024 |
| **Working Status** |  |  |  |  |  |  |
| Working (Full time) | **1.00** |  | **1.00** |  | **1.00** |  |
| Working (Part time) | 1.11 (0.90, 1.38) | 0.329 | 0.91 (0.76, 1.10) | 0.324 | 0.68 (0.56, 0.84) | <0.001 |
| No working/Student | 0.96 (0.79, 1.17) | 0.714 | 0.36 (0.31, 0.43) | <0.001 | 0.27 (0.22, 0.34) | <0.001 |
| **Household factor** |  |  |  |  |  |  |
| **Living Arrangement** |  |  |  |  |  |  |
| Living with Family | **1.00** |  | **1.00** |  | **1.00** |  |
| Living Alone | 1.60 (1.33, 1.93) | <0.001 | 1.67 (1.41, 1.98) | <0.001 | 1.71 (1.40, 2.08) | <0.001 |
| Shared accommodation/Living with Flatmates | 1.02 (0.75, 1.40) | 0.883 | 1.59 (1.23, 2.06) | <0.001 | 2.26 (1.69, 3.02) | <0.001 |
| **Household Size** |  |  |  |  |  |  |
| 1-2 People | **1.00** |  | **1.00** |  | **1.00** |  |
| 3-4 People | 0.76 (0.62, 0.93) | 0.007 | 0.74 (0.61, 0.88) | 0.001 | 0.58 (0.47, 0.70) | <0.001 |
| **Have you been tested for COVID-19?** |  |  |  |  |  |  |
| No | **1.00** |  | **1.00** |  | **1.00** |  |
| Yes, I Tested Negative | 1.57 (1.23, 2.01) | <0.001 | 1.77 (1.42, 2.21) | <0.001 | 1.22 (0.94, 1.57) | 0.136 |
| Yes. I Tested Positive | 1.38 (0.95, 2.01) | 0.095 | 1.42 (1.01, 1.99) | 0.046 | 1.10 (0.75, 1.60) | 0.636 |
| **Compliance with Public health measures** |  |  |  |  |  |  |
| **Have You Enforced Protective Measures Inside Your Home to Protect Yourself and Your Family from COVID 19?** |  |  |  |  |  |  |
| Yes | **1.00** |  | **1.00** |  | **1.00** |  |
| **No** | 1.08 (0.85, 1.38) | 0.540 | 1.18 (0.94, 1.47) | 0.148 | 1.28 (1.00, 1.65) | 0.049 |
| **Are You Currently in Self-Quarantine Since Past Seven Days?** |  |  |  |  |  |  |
| No | **1.00** |  | **1.00** |  | **1.00** |  |
| Yes | 1.28 (1.07, 1.53) | 0.007 | 2.09 (1.75, 2.49) | <0.001 | 1.92 (1.55, 2.38) | <0.001 |
| **Avoid Public Transport** |  |  |  |  |  |  |
| Yes | **1.00** |  | **1.00** |  | **1.00** |  |
| No | 1.08 (0.80, 1.47) | 0.601 | 1.60 (1.20, 2.13) | 0.001 | 4.02 (2.74, 5.91) | <0.001 |
| **Avoid Shaking Hands** |  |  |  |  |  |  |
| Yes | **1.00** |  | **1.00** |  | **1.00** |  |
| No | 0.94 (0.70, 1.27) | 0.700 | 0.68 (0.52, 0.90) | 0.007 | 1.63 (1.15, 2.30) | 0.006 |
| **Wearing Face Mask when going out** |  |  |  |  |  |  |
| Yes | **1.00** |  | **1.00** |  | **1.00** |  |
| No | 1.25 (0.95, 1.64) | 0.107 | 1.15 (0.89, 1.48) | 0.291 | 1.64 (1.19, 2.25) | 0.002 |
| **Avoid Large Gatherings** |  |  |  |  |  |  |
| Yes | **1.00** |  | **1.00** |  | **1.00** |  |
| No | 1.12 (0.86, 1.47) | 0.398 | 1.72 (1.35, 2.19) | <0.001 | 1.18 (0.88, 1.58) | 0.257 |
| **Advocating People About the Health Risk Related to COVID-19** |  |  |  |  |  |  |
| Yes | **1.00** |  | **1.00** |  | **1.00** |  |
| No | 0.55 (0.45, 0.68) | <0.001 | 0.99 (0.81, 1.21) | 0.916 | 0.66 (0.51, 0.84) | 0.001 |
| **Health Condition experienced in previous before the survey** |  |  |  |  |  |  |
| **Fever** |  |  |  |  |  |  |
| No | **1.00** |  | **1.00** |  | **1.00** |  |
| Yes | 0.96 (0.78, 1.17) | 0.663 | 1.11 (0.93, 1.32) | 0.252 | 1.07 (0.88, 1.31) | 0.509 |
| **Chills** |  |  |  |  |  |  |
| No | **1.00** |  | **1.00** |  | **1.00** |  |
| Yes | 1.99 (1.62, 2.44) | <0.001 | 2.07 (1.72, 2.49) | <0.001 | 2.01 (1.65, 2.46) | <0.001 |
| **Headache** |  |  |  |  |  |  |
| No | **1.00** |  | **1.00** |  | **1.00** |  |
| Yes | 1.26 (1.08, 1.48) | 0.004 | 1.63 (1.42, 1.86) | <0.001 | 1.50 (1.29, 1.75) | <0.001 |
| **Cough** |  |  |  |  |  |  |
| No | **1.00** |  | **1.00** |  | **1.00** |  |
| Yes | 1.27 (1.08, 1.50) | 0.004 | 1.79 (1.56, 2.07) | <0.001 | 1.32 (1.12, 1.55) | 0.001 |
| **Breathing Difficulty** |  |  |  |  |  |  |
| No | **1.00** |  | **1.00** |  | **1.00** |  |
| Yes | 2.14 (1.74, 2.62) | <0.001 | 1.73 (1.44, 2.08) | <0.001 | 2.18 (1.79, 2.66) | <0.001 |
| **Dizziness** |  |  |  |  |  |  |
| No | **1.00** |  | **1.00** |  | **1.00** |  |
| Yes | 1.45 (1.20, 1.76) | <0.001 | 1.91 (1.63, 2.25) | <0.001 | 1.50 (1.25, 1.80) | <0.001 |
| **Sore Throat** |  |  |  |  |  |  |
| No | **1.00** |  | **1.00** |  | **1.00** |  |
| Yes | 1.33 (1.11, 1.60) | 0.002 | 1.38 (1.18, 1.61) | <0.001 | 1.39 (1.16, 1.66) | <0.001 |
| Adjusted odd ratios (aOR); 95% confidence intervals (CI) |  |  |  |  |  |  |

## Discussion

This cross-sectional survey assessed the prevalence and associated factors of mental health symptoms related to the COVID-19 pandemic in Bangladesh using an online survey. The study found a high prevalence of mental health symptoms during the pandemic, particularly feeling anxious and depressed while about half of the respondents experienced stress. Respondents who reported these mental health symptoms during the pandemic were more likely to be females, married, have lower education, faced by various factors related to accommodation and living arrangements, and those who experienced COVID-19 related health symptoms. In addition, respondents who were tested for COVID-19, those who traveled via public transport, and people who practiced self-quarantine self-reported a higher prevalence of depression, anxiety, and stress in this study.

The prevalence of mental health symptoms found in this study was higher than previous studies conducted elsewhere during the COVID-19 pandemic including those from the United Kingdom [**20**] and Iran [**21**] where two in five respondents reported some form of depression. Similarly, studies in Asia have shown that people experienced substantial psychological problems during the pandemic [**22, 23**]. The authors attributed factors such as the recommended self-quarantine and isolation measures, employment uncertainty, and the rapid spread of COVID-19 related misinformation [**24**]. A similar high prevalence of depression and anxiety was reported among those with chronic medical problems in Ethiopia during the COVID-19 pandemic [**25**], and in Australia, a high prevalence of psychological distress was reported during the highly infectious equine influenza in 2007 [**26**]. The higher prevalence of mental health symptoms found in this study compared to previous studies may be due to the difference in the study population between studies as well as the socio-cultural differences and the methods used in assessing the mental health symptoms.

Previous epidemiological studies found that women had a higher risk of depression [**27**] than men and were more vulnerable to stress and post-traumatic stress disorder [**28**]. These findings were corroborated in recent studies where the prevalence of anxiety, depression, and stress during the COVID-19 pandemic was significantly higher among women than men [**29, 30, 31**]. Compared with the previous studies the present study used a larger sample size to confirm that women in Bangladesh experienced significantly higher mental health symptoms than men and this could be due to the higher representation of women in various industries like retail, manufacturing, healthcare, and service industries, affected by the current pandemic. These findings are also in agreement with the suggestion that, with the uneven effects in the employment sector, women were more likely to experience psychological and mental health problems when faced with depression, anxiety, and stress [**32**].

Older people had a higher risk of COVID-19 infection and mortality [**33**]., however, the results of existing studies found higher levels of anxiety, depression, and stress among the younger population particularly those aged 21-40years [**11, 34**]. In the present study, age had a significant effect on the participants’ report of mental health symptoms of depression, anxiety, and stress. This may be attributed to the fact that people in this age group are more concerned over the future consequences and economic challenges caused by the pandemic, as they are the key actors working force in society and are, therefore, mostly affected by fear of joblessness and business closures [**35**]. Some researchers have argued that greater anxiety among young people may be related to their greater access to information through social media, which can also cause severe stress [**22, 36**]. It has also been suggested that people become stressed and feel anxious when the information from public health experts are unreliable or was delivered incorrectly and as such could create confusion regarding the practice of self-quarantine or other public health measures put in place to control the spread of a pandemic [**37**].

This study found a significant association between level of education and the mental health symptoms during the pandemic which was consistent with a study from China that was conducted at the initial stage of the COVID-19 outbreak [**38**]. In that study, those with no formal education were more likely to report depression during the epidemic. Other studies have also reported significant associations between the lower level of education, and anxiety and depression levels [**20, 25**]. In contrast to these studies, we found that during the COVID-19 pandemic, respondents in Bangladesh who had a higher level of education reported higher levels of anxiety, depression, and stress [**31, 39**]. It was because 59.75% of respondents of our study had the university level of education in comparison to respondents of the previous studies. Even some recent studies also revealed similar findings as our study did [**40**]

Similar to the findings of this study, some authors in China [**38, 41**] reported higher levels of anxiety among participants who had at least one family member, relative, or friend with COVID-19. Also, those who were separated or widowed/divorced were more likely to experience depression during the pandemic than other respondents which was consistent with a previous study in the USA [**42**] during the pandemic. Before the pandemic, a study in Pakistan revealed that among the older population, those who lived in a nuclear family system were more likely to report depression compared to those who lived in a joint family system [**43**].

During the early outbreak of COVID-19 in Bangladesh, people who had potentially come into contact with the infection were asked to isolate themselves at home or in a dedicated quarantine facility [**1**]. The findings of negative psychological effects of these measures in this study have consisted with other studies which found that lockdown and self-quarantine during the pandemic like SARS, MARS, and COVID-19 had higher levels of post-traumatic stress symptoms, confusion, and anger [**29, 36**]. In our study, more than two-thirds of those who practiced self-quarantine reported mental health issues. Due to the self-quarantine measures and fear related to the spread of COVID-19, other persistent mental health disorders like anxiety, emotional disruption, and exhaustion, depression, anger, irritability, insomnia, and stress can be developed among the population [**44**]. Moreover, the longer the quarantine or self-isolation, the more detrimental these outcomes can become [**45**].

This study had some limitations. First, it was limited in scope. Many of the participants (50%) were from the capital city of Dhaka division, limiting the generalization of our findings to rural regions. Second, the study was carried out during the COVID-19 lockdown period and lacked longitudinal follow-up. The analysis of the periodic state of individuals may not reflect the psychological state, which changes with time and with the alterations in one’s surrounding environment. Because of the increasingly arduous situation and the fear of the second wave, the mental health symptoms of residents could become more severe. Thus, the long-term psychological implications of this population are worth further investigation. Third, due to ethical requirements on anonymity and confidentiality, we were not allowed to collect contact details and personal information from the respondents. As a result, we could not conduct a prospective study that would provide a concrete finding to support the need for a focused public health initiative. Fourth, the study was not able to distinguish between pre-existing mental health symptoms and new symptoms. Fifth, there was an oversampling of a particular network of peers (e.g., students), which may lead to selection bias. However, a large percentage of respondents who were within 40 years old consisted of students. They were exposed to higher mental health symptoms due to the temporary shutdown of educational institutions, disbandment of social gatherings, and the pressure from having to attend classes online [**46**]. Sixth, the self-reported levels of psychological impact, anxiety, depression, and stress may not always align with assessment by mental health professionals. Despite these limitations, this study has several strengths. Further, then providing insight on the actual existing pandemic situation, this study sheds added light on the impact of COVID-19 on the mental health conditions of the educated people in Bangladesh who are the potential future workforce as well.

## Conclusions

In this survey study conducted in Bangladesh, respondents reported high rates of symptoms of depression, anxiety, and distress while non-compliance with public health measures increased the risk of mental health outcomes. Protecting the Bangladesh population is an important component of public health measures for addressing the COVID-19 epidemic. Special interventions to promote mental wellbeing in Bangladesh communities exposed to COVID-19 need to be immediately implemented, with women, married people, the less educated people, and those that were tested for COVID-19 requiring particular attention. To further close the gap in the relationship and improve the mental health wellbeing of the Bangladeshi people, alternative ways of communication such as the use of internet video calls [**47**] should be promoted during similar situations.

## Data Availability

This is necessary that we stored the data in any repository. However, our data are included in the manuscript and raw data can be released on reasonable requests.

---

**Supplementary Table 1:** Sample of survey items and their test of reliability.

| Did not apply to me<br>at all (0) | Applied to me to some degree or<br>some of the time (1) | Applied to me to a considerable degree or<br>most of the time (2) | Applied to me very much or most<br>of the time (3) |
| --- | --- | --- | --- |
| Depression subscale |  |  | Cronbach's Alpha |
| 3. I couldn't seem to experience any positive feeling at all |  |  | 0.7618 |
| 5. I found it difficult to work up the initiative to do things |  |  | 0.7661 |
| 10. I felt that I had nothing to look forward to |  |  | 0.7594 |
| 13. I felt downhearted and blue |  |  | 0.7690 |
| 16. I was unable to become enthusiastic about anything |  |  | 0.7650 |
| 17. I felt I wasn't worth much as a person |  |  | 0.7330 |
| 21. I felt that life was meaningless |  |  | 0.7633 |
| Anxiety subscale |  |  | Cronbach's Alpha |
| 2. I was aware of dryness of my mouth |  |  | 0.6644 |
| 4. I experienced breathing difficulty (e.g. excessively rapid breathing, breathlessness in the absence of physical exertion) |  |  | 0.6768 |
| 7. I experienced trembling (e.g. in the hands) |  |  | 0.7720 |
| 9. I was worried about situations in which I might panic and make a fool of myself |  |  | 0.6668 |
| 15. I felt I was close to panic |  |  | 0.6648 |
| 19. I was aware of the action of my heart in the absence of physical exertion (e.g. sense of heart rate increase, heart missing a beat) |  |  | 0.6513 |
| 20. I felt scared without any good reason |  |  | 0.6511 |
| Stress subscale |  |  | Cronbach's Alpha |
| 1. I found it hard to wind down |  |  | 0.8100 |
| 6. I tended to over-react to situations |  |  | 0.7894 |
| 8. I felt that I was using a lot of nervous energy |  |  | 0.7776 |
| 11. I found myself getting agitated |  |  | 0.7877 |
| 12. I found it difficult to relax |  |  | 0.7775 |
| 14. I was intolerant of anything that kept me from getting on with what I was doing |  |  | 0.7822 |
| 18. I felt that I was rather touchy |  |  | 0.7823 |

**Supplementary Table 2**

**A survey questionnaire on Mental health and psychological impact of the coronavirus disease 2019 (COVID-19) outbreak: A cross-sectional study on Bangladeshi People**

## Socio-Demographic Profile of the Respondent

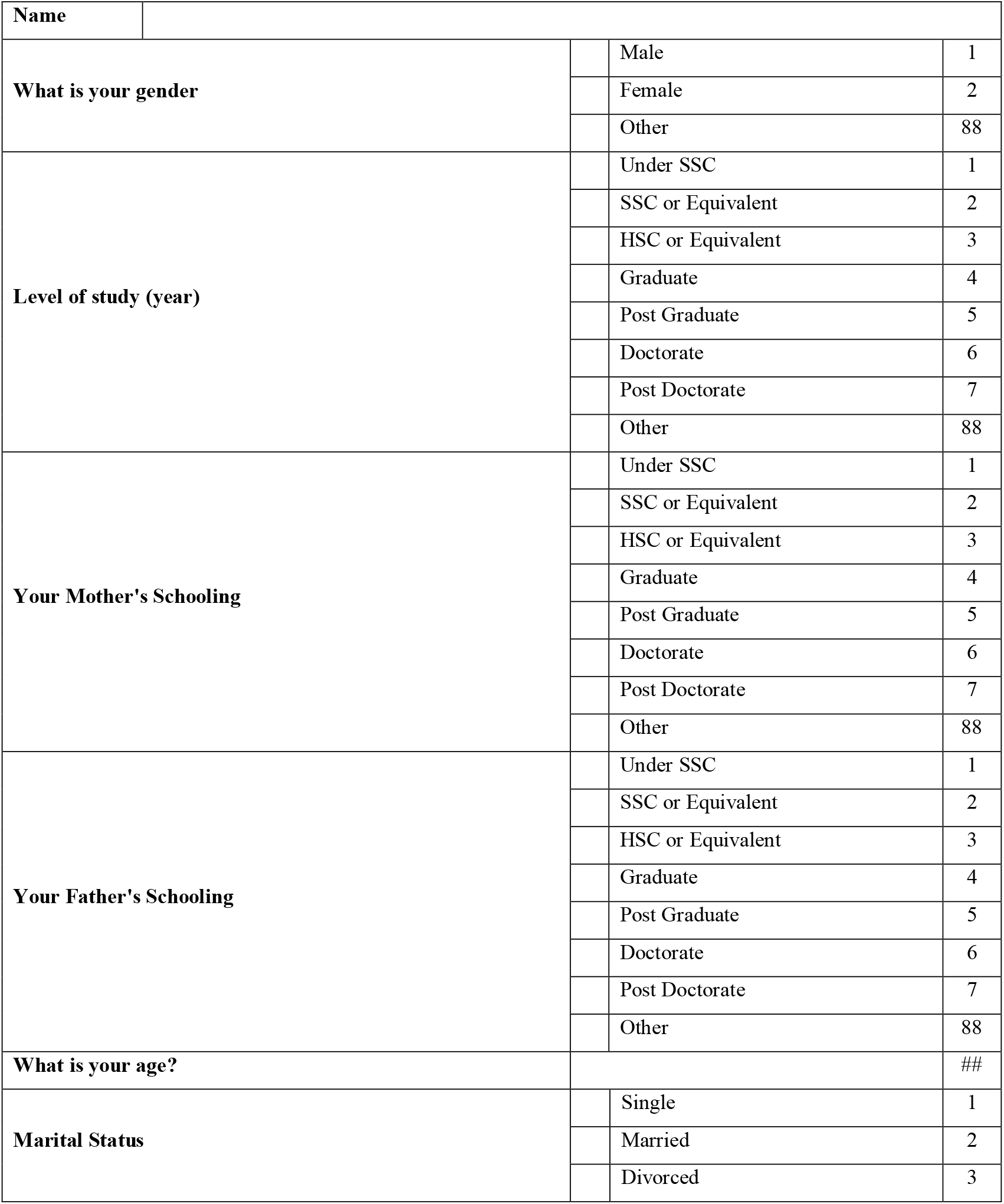
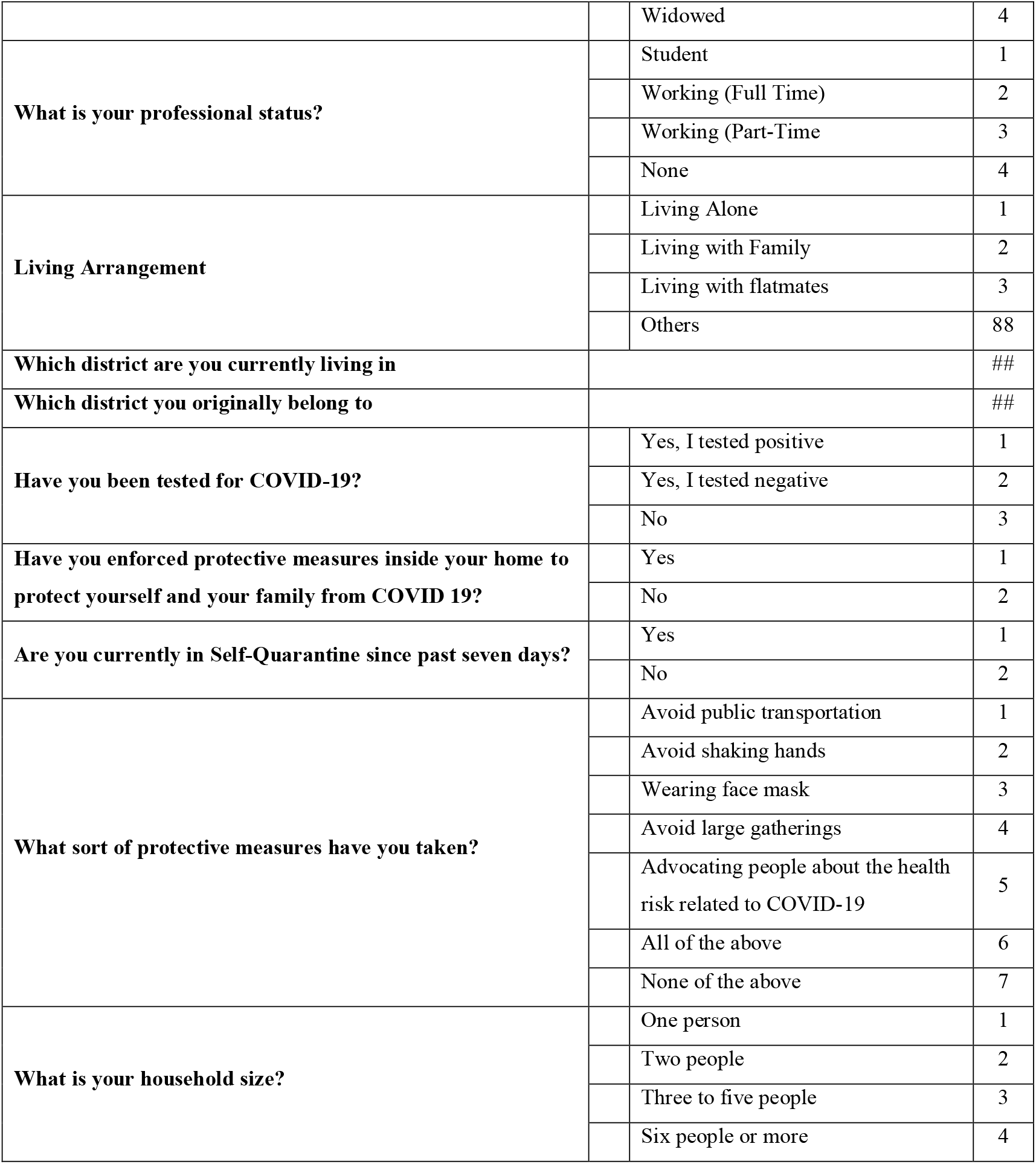

Socio-Demographic Profile of the Respondent:
| Name |  |  |  |
| --- | --- | --- | --- |
| What is your gender |  | Male | 1 |
|  |  | Female | 2 |
|  |  | Other | 88 |
| Level of study (year) |  | Under SSC | 1 |
|  |  | SSC or Equivalent | 2 |
|  |  | HSC or Equivalent | 3 |
|  |  | Graduate | 4 |
|  |  | Post Graduate | 5 |
|  |  | Doctorate | 6 |
|  |  | Post Doctorate | 7 |
|  |  | Other | 88 |
| Your Mother's Schooling |  | Under SSC | 1 |
|  |  | SSC or Equivalent | 2 |
|  |  | HSC or Equivalent | 3 |
|  |  | Graduate | 4 |
|  |  | Post Graduate | 5 |
|  |  | Doctorate | 6 |
|  |  | Post Doctorate | 7 |
|  |  | Other | 88 |
| Your Father's Schooling |  | Under SSC | 1 |
|  |  | SSC or Equivalent | 2 |
|  |  | HSC or Equivalent | 3 |
|  |  | Graduate | 4 |
|  |  | Post Graduate | 5 |
|  |  | Doctorate | 6 |
|  |  | Post Doctorate | 7 |
|  |  | Other | 88 |
| What is your age? |  |  | ## |
| Marital Status |  | Single | 1 |
|  |  | Married | 2 |
|  |  | Divorced | 3 |

|  |  |  |
| --- | --- | --- |
|  | Widowed | 4 |
| **What is your professional status?** | Student | 1 |
|  | Working (Full Time) | 2 |
|  | Working (Part-Time) | 3 |
|  | None | 4 |
| **Living Arrangement** | Living Alone | 1 |
|  | Living with Family | 2 |
|  | Living with flatmates | 3 |
|  | Others | 88 |
| **Which district are you currently living in** |  | ## |
| **Which district you originally belong to** |  | ## |
| **Have you been tested for COVID-19?** | Yes, I tested positive | 1 |
|  | Yes, I tested negative | 2 |
|  | No | 3 |
| **Have you enforced protective measures inside your home to protect yourself and your family from COVID 19?** | Yes | 1 |
|  | No | 2 |
| **Are you currently in Self-Quarantine since past seven days?** | Yes | 1 |
|  | No | 2 |
| **What sort of protective measures have you taken?** | Avoid public transportation | 1 |
|  | Avoid shaking hands | 2 |
|  | Wearing face mask | 3 |
|  | Avoid large gatherings | 4 |
|  | Advocating people about the health risk related to COVID-19 | 5 |
|  | All of the above | 6 |
|  | None of the above | 7 |
| **What is your household size?** | One person | 1 |
|  | Two people | 2 |
|  | Three to five people | 3 |
|  | Six people or more | 4 |

## Physical Health Status

(Please respond on your physical health since the past two weeks)

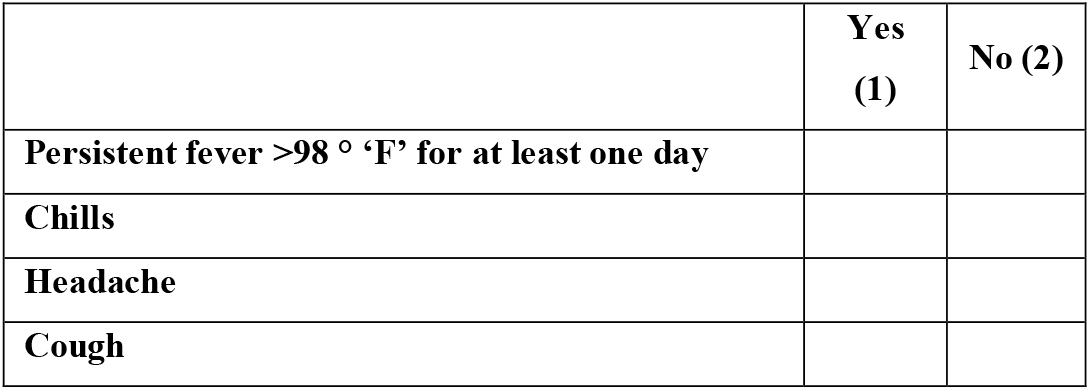
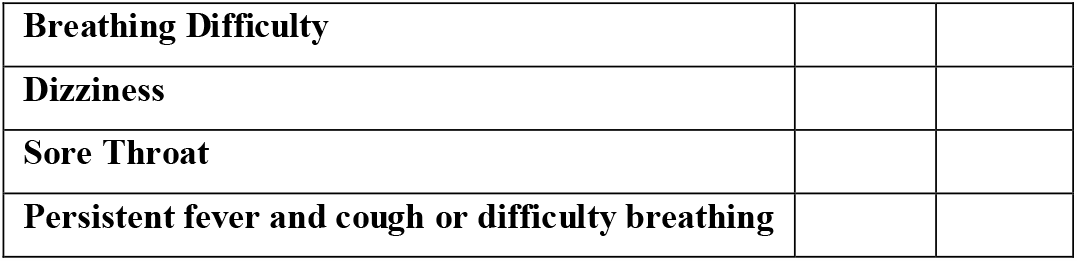

## Mental Health Impact of COVID 19

Please read each statement and circle a number 1, 2, 3 or 4 which shows how much the statement has applied in the past week to you. There are no correct answers or incorrect answers.

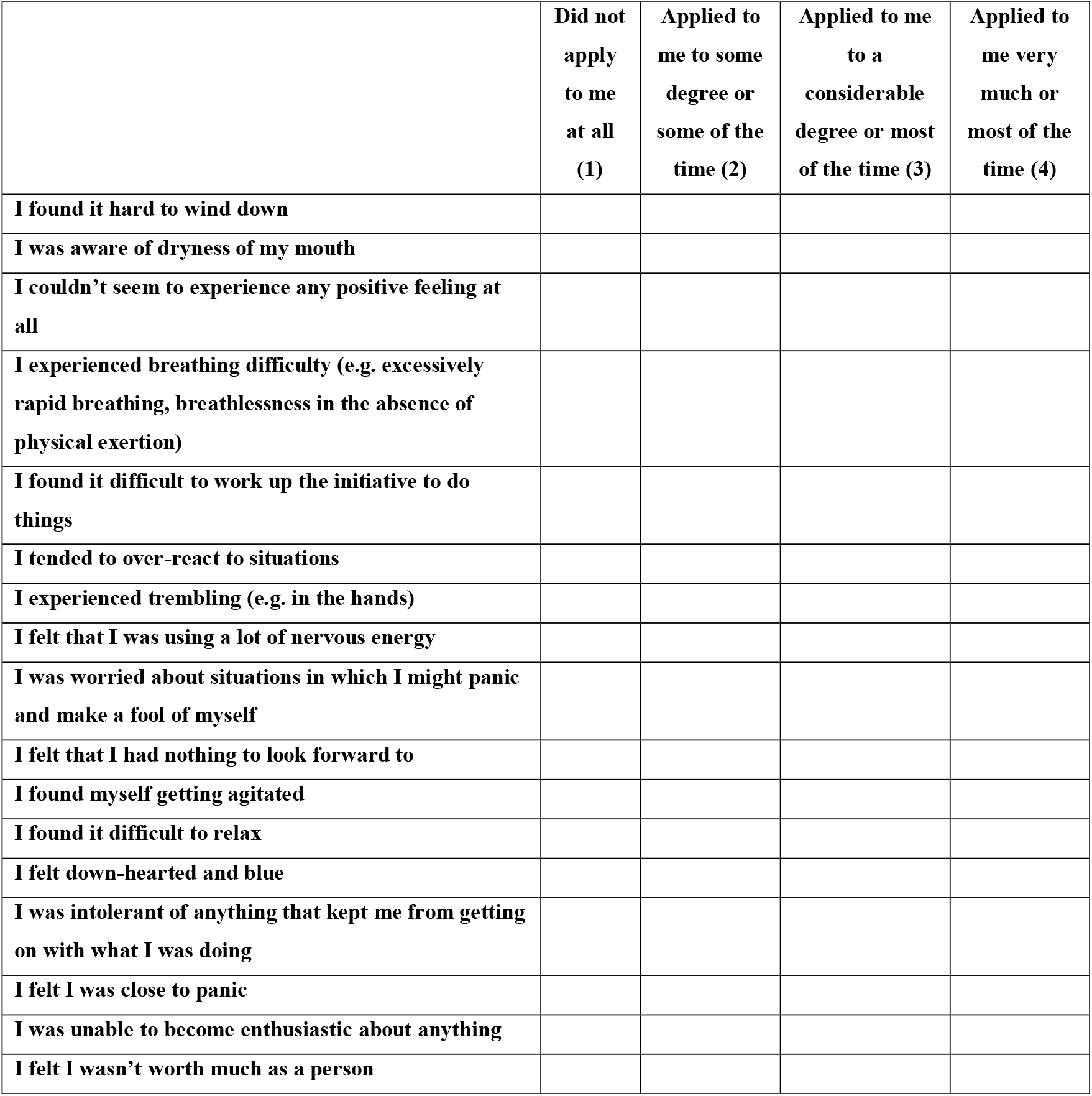
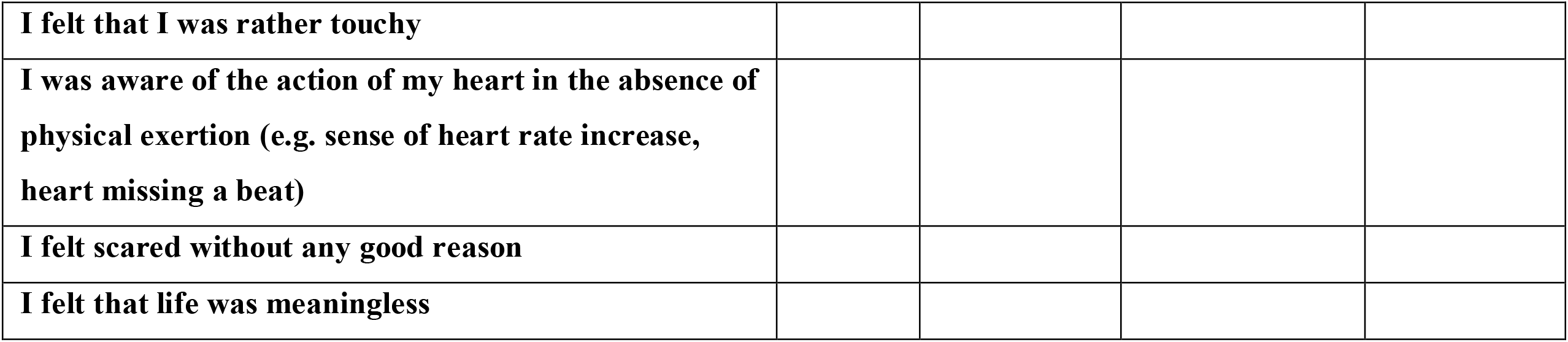

Thank you for your participation

